# Epidemiology of malignant hyperthermia in the UK 1988-2025: implications for prevalence, mode of inheritance, relative risk associated with *RYR1* genotypes and *in vitro* contracture test phenotype

**DOI:** 10.64898/2026.03.05.26347692

**Authors:** Essameldeen M Aboelsaod, Catherine Daly, Nickla Fisher, Sarah Hobson, Huw Garland, Pawan K Gupta, Jonathan G Bilmen, Sarah Shepherd, Rachel L Robinson, Marie-Anne Shaw, Philip M Hopkins

**Author notes:** Corresponding author: PM Hopkins.

## Abstract

**Background:** There is disparity between the incidence of malignant hyperthermia (MH) reactions and the prevalence of variants in the *RYR1* gene associated with susceptibility to MH (where susceptibility is determined by *in vitro* contracture tests). Our aims were to use clinical and genetic data from the UK to explain this disparity and to examine if these data are consistent with the clinical risk of MH being inherited as an autosomal dominant trait.

**Methods:** Clinical MH and genotyping data were extracted from the UK MH registry. The numbers of general anaesthetics delivered in the UK were estimated from national surveys and reports, with population data obtained from government statistics. The prevalence of *RYR1* variants in the UK population was estimated using UK Biobank data. The incidence of MH reactions 1988-93 was used to estimate the prevalence of the clinical risk of MH in the UK. Bayesian modelling, calibrated against actual data, was used to evaluate the likely mode of inheritance of the clinical risk of MH and the relative risk of clinical MH associated with different *RYR1* variants.

**Results:** The probability of index cases developing MH with each general anaesthetic can be expressed as a constant hazard of 0.46 (95% CI 0.42 – 0.50, n=375). We used peak incidence data (1988-93) to estimate the prevalence of the risk of MH as 1 in 44,000 (95% credibility interval, 1 in 40,000 to 1 in 48,000). The incidence of MH has declined over the past 22 years but the rate of decline is inconsistent with autosomal dominant inheritance (*P* < 10^-10^). The risk of MH varied by up to 150-fold between carriers of 28 recurrent *RYR1* variants.

**Conclusion:** These findings support a threshold inheritance model for clinical MH and have implications for diagnostics, both genotyping and *in vitro* contracture test phenotyping.

---

Malignant hyperthermia (MH) is an adverse reaction to potent inhalation anaesthetics or succinylcholine caused by dysregulation of skeletal muscle calcium ion homeostasis.^1^ Its cardinal clinical features are a result of intense and sustained stimulation of skeletal muscle metabolism and include increased CO_2_ production, O_2_ consumption, heart rate and body temperature. Other features may include muscle rigidity, rhabdomyolysis and coagulopathy.^2,3^

MH has a genetic basis, with inheritance of the risk of MH described as autosomal dominant with variable penetrance in the initial report of the condition.^4^ Three genes, *RYR1*, *CACNA1S* and *STAC3*, have been implicated as contributing to the risk of MH with *RYR1* the most commonly implicated, but approximately 25% of MH families do not harbour variants in any of these genes.^5^ Research into the genetics of the risk of MH is challenging because those carrying the risk typically have no clinical phenotype unless exposed to general anaesthesia involving the triggering drugs: even then, those who have developed MH reactions often report uneventful previous anaesthesia.

The surrogate phenotype used in many genetic studies is the MH susceptibility phenotype, which is defined by the results of *in vitro* contracture tests (IVCTs).^6^ IVCTs, in which freshly excised strips of quadriceps muscle are exposed under standardised laboratory conditions separately to halothane and caffeine, have been the “gold standard” diagnostic test for the risk of MH since they were described more than 50 years ago.^7,8^ The IVCT described by the European MH Group has point estimates for sensitivity of 100% and specificity of 94%.^9,10^ While segregation of the IVCT phenotype with genetic markers within affected families enabled identification of the *RYR1* and *CACNA1S* loci,^11–13^ family cascade testing using the IVCT has assumed a classical Mendelian autosomal dominant inheritance. If this assumption is false, analyses of the inheritance pattern of the risk of MH that depend on family studies using the IVCT phenotype will be biased. A further assumption that may have confounded thinking in the field is that all those with the MH susceptibility (IVCT) phenotype carry the same all-or-none risk of developing a clinical MH reaction. These assumptions have also been made by the ClinGen variant curation expert panel when designing criteria to determine the pathogenicity of *RYR1* variants in relation to the risk of MH.^14,15^

Data challenging monogenic inheritance of the risk of MH have been available for more than 30 years. They include the presence of discordance between *RYR1* genotype and IVCT phenotype within affected families,^16–18^ with examples of both genotype-positive/phenotype-negative and genotype-negative/phenotype-positive individuals.^19,5^ There is also a high incidence of affected families in which more than one potentially pathogenic variant in *RYR1*, *CACNA1S*, or both is present.^5^

In this study, we aim to estimate the proportion of the UK population who are at risk of developing a clinical MH reaction and contrast this to the prevalence of *RYR1* variants associated with that risk. Based on these estimates we will model the annual number of expected cases of MH, calibrated to actual data, and evaluate the plausibility of clinical risk being inherited in an autosomal dominant fashion. We further aim to determine if the risk of developing a clinical MH reaction varies according to the specific *RYR1* variant present in the index case and whether that risk is associated with the strength of responses to halothane and caffeine measured in the IVCT.

## Methods

### Patients and genetic analyses

De-identified MH patient data were extracted from the database of the Leeds MH Investigation Unit, which provides a national MH diagnostic service for the whole of the United Kingdom. We included data only from families where the index case presented with a suspected MH reaction and excluded data where the index case did not have an adverse anaesthetic event, presenting alternatively with, for example, exertional rhabdomyolysis or congenital myopathy. For index cases we included the number of uneventful anaesthetics they had received before the index event, the year of the index event and the year their diagnosis was confirmed (by IVCT or genotyping). For family members, we included the number of uneventful anaesthetics they had received up to the date of the index event and the year their diagnosis was confirmed. IVCT phenotyping and *RYR1* genotyping were as described in Miller et al.^5^ All patients gave written informed consent and the research was approved by Leeds (East) Research Ethics Committee or its predecessors: Leeds Teaching Hospitals NHS Trust Clinical Research (Ethics) Committee (East) and Leeds Health Authority / St James’s and Seacroft University Hospitals Clinical Research (Ethics) Committee.

### Prevalence of the risk of developing a clinical MH reaction in the UK population

This requires an estimate of the number of MH reactions (numerator), the number of general anaesthetics administered using MH triggering drugs (denominator) and the probability that an individual who has the risk of developing a clinical reaction reacts with each exposure to general anaesthesia (correction factor). For these analyses, we excluded index cases that had not occurred in the UK.

### Number of MH reactions

The number of patients suspected of having a possible MH reaction, as well as the number with confirmed (by IVCT) MH reactions, has varied over the past 55 years. However, it is reasonable to assume that the proportion of the UK population at risk of developing a reaction is stable over time, with the variability accounted for by clinical acumen, reporting compliance, changes in anaesthetic technique and avoidance of exposure to trigger agents following emergence of a family history of a possible MH reaction. Our estimate is therefore based on the six year period with the highest number of confirmed cases (1988-93) in the UK. Our experience is that, within a national health service with a single diagnostic unit for MH, the capture rate, including patients with delayed referral and/or diagnosis, is high: our point estimate therefore assumes that 95% of cases in this six year period were reported. A sensitivity analysis will evaluate low and high capture rates of 90% and 100% respectively.

### Number of general anaesthetics administered in the UK

Surgical procedure data were not routinely collated in the UK in 1988-93 but the National Confidential Enquiry into Perioperative Deaths (NCEPOD) requested the Department of Health to provide such data for the calendar year 1989.^20^ These data were for England only but we assume procedure rates were similar across the remaining 16% of the UK population. The Department of Health data excluded obstetric procedures^21–23^ and those procedures carried out in the private health sector^24^ and adjustments were made to reflect these omissions. Adjustments were not made for community dental general anaesthesia because these brief exposures to inhalation anaesthesia, some using nitrous oxide only, have not led to confirmed MH cases. The proportion of surgical procedures carried out under general anaesthesia was estimated to be 80% in 2014^25^ but 25 years earlier UK practice was quite different (less frequent use of local and regional anaesthesia). We therefore estimated that 90% of non-obstetric surgical procedures were carried out under general anaesthesia and that 95% of general anaesthetics included exposure to MH triggering drugs. To calculate the proportion of the population receiving general anaesthesia each year over the 6 year period we used UK mid-year estimates provided by the Office for National Statistics.^26^ Sensitivity analysis will evaluate: low (85%) and high (95%) estimates of the proportion of non-obstetric surgical procedures carried out under general anaesthesia; low (90%) and high (100%) estimates of the proportion of general anaesthetics that used MH triggering drugs: low (10%), midpoint (12.5%) and high (15%) estimates for the proportion of elective surgical procedures carried out in the private health sector.

### Probability that an individual who has the risk of developing a clinical reaction reacts with each exposure to general anaesthesia

We searched the MH database for index cases with a complete anaesthetic history and collated how many general anaesthesia exposures each index case had before they developed an MH reaction. We tested two hypotheses:

1. All index cases have a similar risk of reacting, so that the risk of reacting with each exposure is constant and equal to the overall proportion of reactions to exposures (to the nearest 10%);
2. Some index cases are more resilient than others, so that the risk of reacting with each exposure successively reduces as less resilient patients are identified by the earlier exposures.

### “Stress-testing” estimates using actual data

While the assumptions used in our estimates are based on the best available evidence and the considerable combined experience of the authors, we can evaluate the rigour of the estimates by using them to model the number of new MH cases presenting each year in the UK from 1988 – 2025, for which we have actual data. We will factor-in a reduction in the proportion of the population receiving general anaesthesia each year (to approximately 5%, based on National Audit Project activity surveys^27,28^) and the reduction in use of MH-triggering anaesthesia (reduced use of succinylcholine and increased use of total intravenous anaesthesia, TIVA) to an estimated current use of 80% of general anaesthetics. The pattern of annual incidence of MH cases will be influenced by the number of related individuals among those at risk, i.e., the inheritance pattern of the risk of MH. We therefore conducted initial Bayesian modelling to estimate the mean number of affected individuals (those at risk of developing a reaction) per family.

### Prevalence of *RYR1* variants in the UK population

To estimate the prevalence of *RYR1* variants in the UK population we used minor allele frequency (MAF) data made freely available by the UK Biobank (https://afb.ukbiobank.ac.uk/gene/RYR1, accessed 10/03/2025). The UK Biobank obtained the MAF data by genome sequencing approximately 490,000 UK volunteers. As the expected allele count for rarer variants was low, we used prevalence from the non-UK Biobank subset of the non-Finnish European cohort of the gnomAD 4.1 dataset (https://gnomad.broadinstitute.org/gene/ENSG00000196218?dataset=gnomad_r4, accessed 15/01/2026) to corroborate the UK Biobank data.

### IVCT data

We used the results of the static halothane and static caffeine IVCTs from 502 members of 203 families in which one of 22 of the most prevalent non-synonymous *RYR1* variants in our UK MH cohort had been found. To quantify the responses in each test we used the maximum contracture (g force) during application of either 2% (v/v) halothane (static halothane test) or 2 mM caffeine (static caffeine test). These data have been previously published by Carpenter et al.^30^

### Data handling and statistical analyses

Data were either imported, or entered directly, into Excel spreadsheets (v16.62 for Mac, Microsoft Corporation).

Evaluation of the hypotheses for the risk of index cases developing an MH reaction with each general anaesthesia exposure first used a constant hazard survival model with Wilson’s method for the calculation of 95% confidence intervals. The alternative hypothesis was tested using a logistic trend model with each risk set as a binomial.

Estimates with 95% credibility intervals for the prevalence of individuals at risk of developing clinical MH (reactors), probabilistic modelling of the number of reactors in each MH family and modelling of the number of MH reactions per year 1988-2025 with 95% credibility intervals used Bayesian Monte Carlo simulations with 2,000-50,000 trials.

To compare the risk of clinical MH reactions for carriers of each *RYR1* variant over the past 55 years the number of MH events was based on the number of independent families in which each variant has been found. The “population” of carriers with each variant was the estimated average number of UK individuals harbouring the variant and this was extrapolated from twice the prevalence of the variant multiplied by 62,000,000 (average UK population between 1970 and 2025). Wilson 95% CIs of binomial proportions were calculated for allele frequencies. To assess if there were any differences in risk of MH reactions between variants we first used Poisson regression to do a likelihood-ratio test. We then did pairwise variant comparisons of likelihood ratios with *P* values corrected using Benjamini–Hochberg false discovery rate (FDR). Relative risks are displayed using a Forest plot.

The association between relative risk of variants and IVCT responses was evaluated using Spearman’s rank correlation coefficient. A *P* value < 0.05 was inferred to be statistically significant, with a Bonferroni correction made for tests involving one of the two IVCT tests (*P* < 0.025).

Statistical analyses were done in Python v3.13 using the following packages: pandas, numPy, math, scipi.stats, statsmodels.api, statsmodels.formula.api, itertools.combinations and os packages, with plots constructed using matplotlib. The Python code was written using ChatGPT 5.1 or 5.2.

Sample sizes were based on convenience samples.

## Results

### Number of MH reactions in the UK 1988-93

There was a total of 150 suspected MH reactions reported to the UK MH Unit and confirmed using IVCT in the 6 year period. The full series of referral and reaction numbers for 1988-2025 is shown in Supplementary Table S1.

### Number of general anaesthetics administered in the UK in 1989

In response to the request from NCEPOD,^20^ the Department of Health identified 2,783,172 non-obstetric surgical procedures conducted in NHS hospitals in England in 1989, from which we can extrapolate that there were 3,313,300 such procedures carried out in the UK. With 11-12% of the 790,000 births in the UK in 1989 being Caesarean deliveries we estimate 50,000 obstetric general anaesthetics.^21–3^ We further assume that 12.5% of the elective surgical workload was done in the private health sector,^24^ 80% of the NHS non-obstetric surgical workload was elective and 90% of the total non-obstetric surgical procedures were carried out under general anaesthesia to derive our point estimate of the number of general anaesthetics in the UK at 3.37 million, or 5.9% of the 57.1 million population. The sensitivity analysis provided low and high estimates of 3.12 million general anaesthetics (5.4% of the population) and 3.64 million general anaesthetics (6.4% of the population) respectively.

### Incidence of malignant hyperthermia reactions in the UK, 1988-93

Based on a mean UK population of 57.25 million, 5.9% of the population receiving general anaesthesia each year, a mean of 25 cases of MH reported each year, reporting completeness (ascertainment) of 95%, our point estimate for the incidence of MH is 7.8 cases per million (1 in 128,000) general anaesthetics. Bayesian Monte Carlo modelled credibility intervals (using low and high estimates for ascertainment and population annual general anaesthesia exposure rate of 0.9 and 1.0, and 0.054 and 0.064 respectively) yielded a 95% credibility interval for the incidence of MH of 7.2 – 8.5 cases per million (1 in 118,000 to 1 in 138,000) general anaesthetics.

### Probability that an individual who has the risk of developing a clinical reaction reacts with each exposure to general anaesthesia

We identified 375 unrelated MH index cases for whom we had a complete anaesthetic history, of which 371 had their MH reaction during their 1^st^ – 6^th^ exposures to general anaesthesia (Table 1). Three of the remaining cases reacted during their 8^th^ exposure, while the final patient reacted only on their 12^th^ exposure to general anaesthesia. The pattern of risk of reaction with each exposure was similar between the 247 male and 128 female index cases (Supplementary Table S2).

**Table 1.**
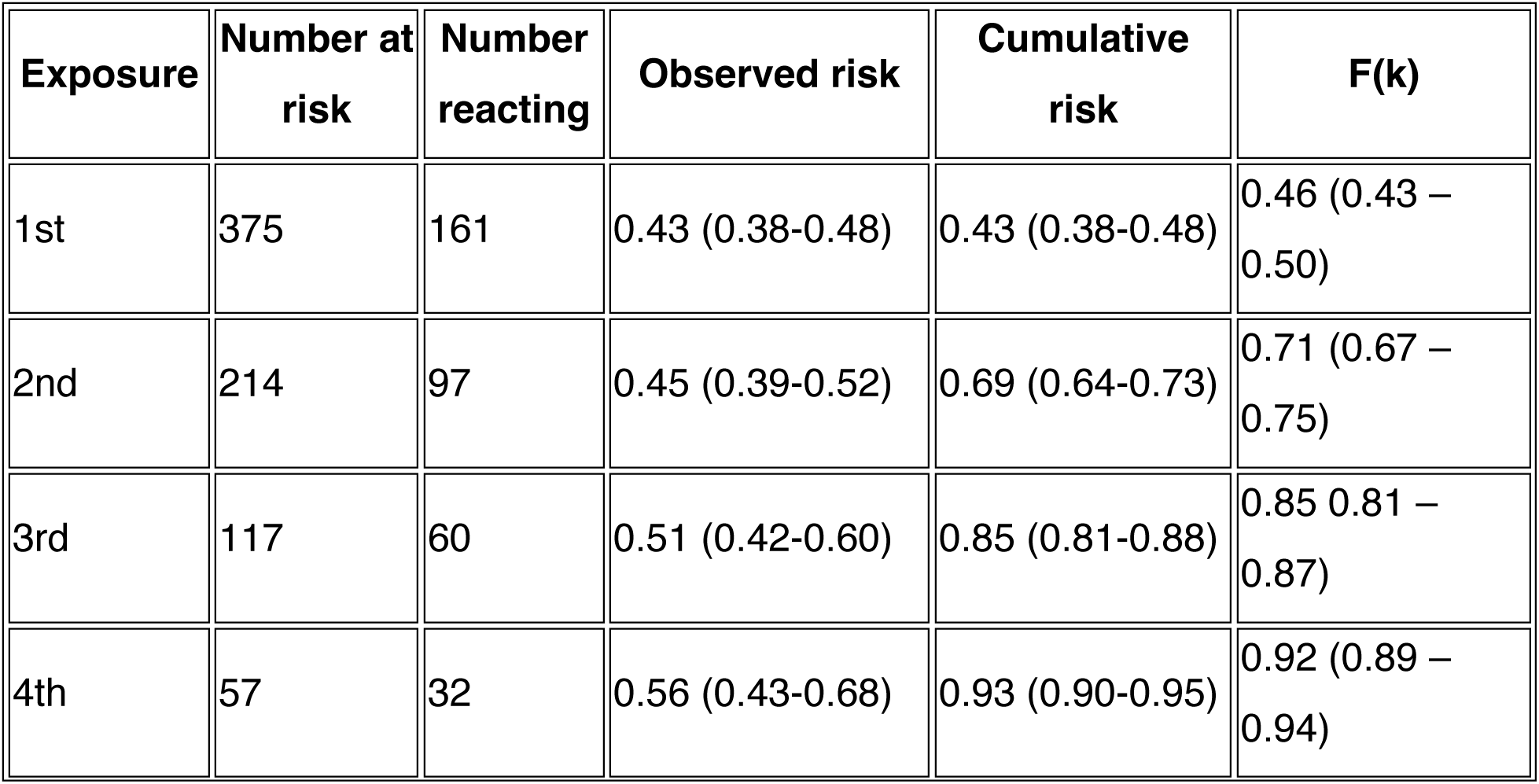
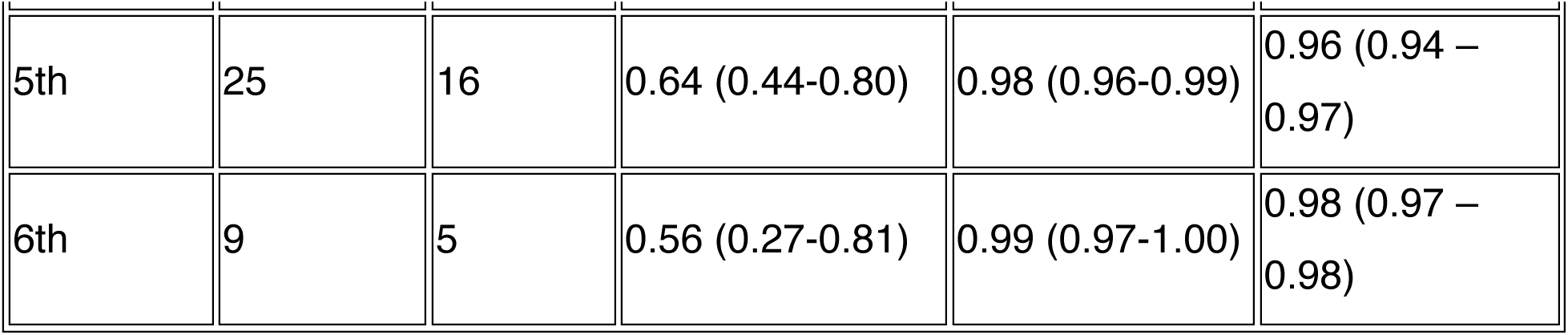
The number of MH probands who reacted with each exposure to general anaesthesia. Observed and cumulative risk is presented as point estimate and 95% binomial CIs. F(k) is the predicted and 95% CI cumulative risk derived from a constant hazard risk model.

For the constant hazard analysis we hypothesise that the risk at each exposure is the same as the overall rate of exposure (375 reactions from 809 exposures) of 0.46 (95% CI 0.42 – 0.50). Table 1 shows the modelled cumulative risk (and 95% CI) of reacting with successive exposures along with the observed cumulative risk, which falls within the modelled plausible range at each exposure.

To test the alternative hypothesis, that the risk of reacting reduces with each exposure (more resilient patients are less likely to react with each exposure), we applied a logistic trend model (a generalised linear model on the logit function). The estimated β coefficient (slope) was 0.172 with 95% CI 0.052 to 0.293, indicating that decreasing risk with successive exposures (in which case β would be < 0) was not supported by the data (*P* = 0.0025, one-tailed).

### Prevalence of the risk of developing a clinical MH reaction in the UK population

Re-running our Bayesian Monte Carlo model for the incidence of MH reactions during the 6 year peak period for MH cases in the UK, but adding triangulated estimates of the proportion of general anaesthetics at that time that involved MH triggering drugs (low 0.9, mid 0.95, high 1.0) and the probability of an individual at risk developing a reaction with each exposure to general anaesthesia (low 0.42, mid 0.46, high 0.50), we can derive estimates (with 95% credibility intervals) of the prevalence of individuals at risk to be 1 in 56,000 (1 in 50,000 to 1 in 62,000) of the UK population. However, prior to 1988, 265 unrelated individuals within the UK had been identified to be at risk of developing MH based on a clinical history of a suspected reaction and a positive IVCT. If we assume that these individuals were not at risk of exposure to triggering agents after their diagnosis, they should be included in an estimate (95% credibility interval) of the prevalence of those capable of developing an MH reaction in the UK of 1 in 44,000 (1 in 40,000 to 1 in 48,000).

### Use of these estimates to predict UK MH cases 1988-2025 and comparison with actual data

In modelling predicted MH cases since 1988, it is necessary to consider changes in the population, rate of general anaesthesia exposure of the UK population, proportion of general anaesthetics involving triggering anaesthetics, ascertainment of MH cases and censoring of those diagnosed with an increased risk of developing MH (and therefore at low risk of exposure to triggering anaesthetics). We assumed a consistent rate of fall of general anaesthesia rate from 5.9% to 5% of the population per annum (the latter based on National Audit Project surveys^27,28^) and a biphasic reduction in use of MH trigger drugs, with the first a relatively rapid decline in the use of succinylcholine from the mid-1990s and a more recent slower decline attributed to increasing use of TIVA. We combined these two factors to input triangulated estimates of exposure decline as follows: fast from 1993–2002 (low 0.10, mid 0.15, high 0.20 %/yr) and slow from 2003-2025 (low 0.04, mid 0.06, high 0.08 %/yr). Examination of our database also indicated that a non-constant rate of ascertainment of cases was appropriate. Our data indicate that ascertainment of cases that occurred more than 15 years ago is high but falls progressively with more recent reactions. We therefore used the same constant triangulated values for ascertainment for reactions that occurred between 1988 and 2010 (low 0.9, mid 0.95, high 1.0) but values that each declined by 0.05 per annum from 2011-2025.

Censoring of cases for 1988 needed to account for those identified before 1988 but censoring for each subsequent year also needed to include the number of confirmed cases referred the previous year (Supplementary Table S1 shows the number of referred cases each year 1988-2025).

However, censoring will take place when family members have been identified as having a MH reaction, so the number censored will depend on the average number of individuals within each family who have the potential to develop an MH reaction (k). If the potential to react clinically is inherited as an autosomal dominant trait, k would be expected to be at least 3 for a compact 3 generation family where the average fertility rate is 2. With increasing family size, k increases further above 3. We therefore next set out to determine k using probabilistic modelling based on actual MH cases from 1988-2025. This modelling, based on an effective posterior resampling size of 50,000 gave median (95% credibility interval) values of 1.18 (1.01-1.43), with the chance of k > 2 having *P* < 10^-10^. In order to check that this estimate of k was not dependent on a too-narrow range of estimates of the prevalence of MH, we re-ran the probabilistic model with high to low estimated prevalence of 1 in 20,000 to 1 in 200,000. This gave median (95% credibility interval) values for k of 1.17 (1.00-1.60), with the chance of k ≥ 2.0 having *P* = 0.0004, chance of k ≥ 2.5 having *P* < 10⁻⁶ and chance of k ≥ 3.0 having *P* < 10^-10^.

Using triangulated values of k (low 1.01, mid 1.18, high 1.43) we then modelled predicted number of reactions from 1988-2025. Figure 1 shows the point estimate with 95% credibility intervals along with the observed number of cases.

**Figure 1.**
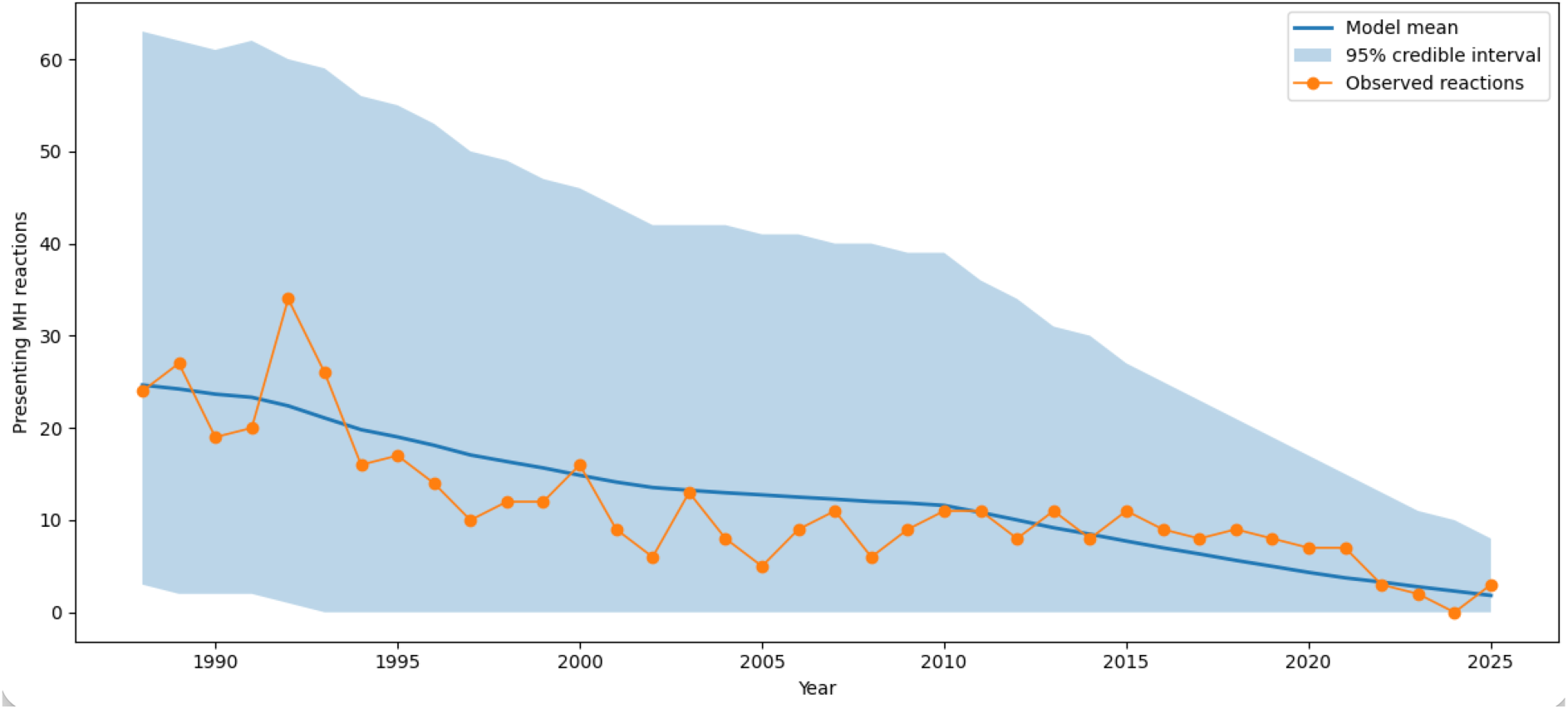
Comparison of actual (observed reactions) and predicted malignant hyperthermia (MH) reactions in the UK 1988-2025. Prediction was based on Bayesian Monte Carlo modelling using 8,000 runs to ensure model stability.

### Relative risk of developing a clinical MH reaction according to *RYR1* genotype

We evaluated 28 *RYR1* variants that have each been found in 5 or more UK MH families. Supplementary table S3 provides the details of theses variants, their prevalence in the UK Biobank database and the non-UK Biobank subset of the European non-Finnish cohort of the gnomAD v4.1.0 database. The 95% CIs of the estimates of the prevalence of each variant overlapped between the two cohorts, providing reassurance that the UK Biobank estimates are reasonable. The p.Arg2435His and p.Val4234Leu variants were not present in the UK Biobank cohort making it difficult to estimate the number of UK carriers. However, as they have been found in 10 and 6 independent UK MH families respectively, it seems likely that carriers are at high risk of developing MH.

Using a Poisson generalised linear model we found the likelihood ratio statistic for a difference in MH risk among the remaining 26 variants to be 538 (25 df, *P* = 1 x 10^-97^). Pairwise comparisons demonstrated many statistically significant differences in risk of MH between variants (supplementary table S4). The relative risk of developing MH with each variant present and its 95% CI is shown in Figure 2. Even excluding p.Arg2435His and p.Val4234Leu, because of the uncertainty in the estimate of their prevalence there was a > 125-fold difference in the relative risk of a clinical MH reaction for carriers of the high risk variant p.Arg163Cys and the variants with the lowest risk (p.Ser1728Phe and p.Pro4973Leu).

**Figure 2.**
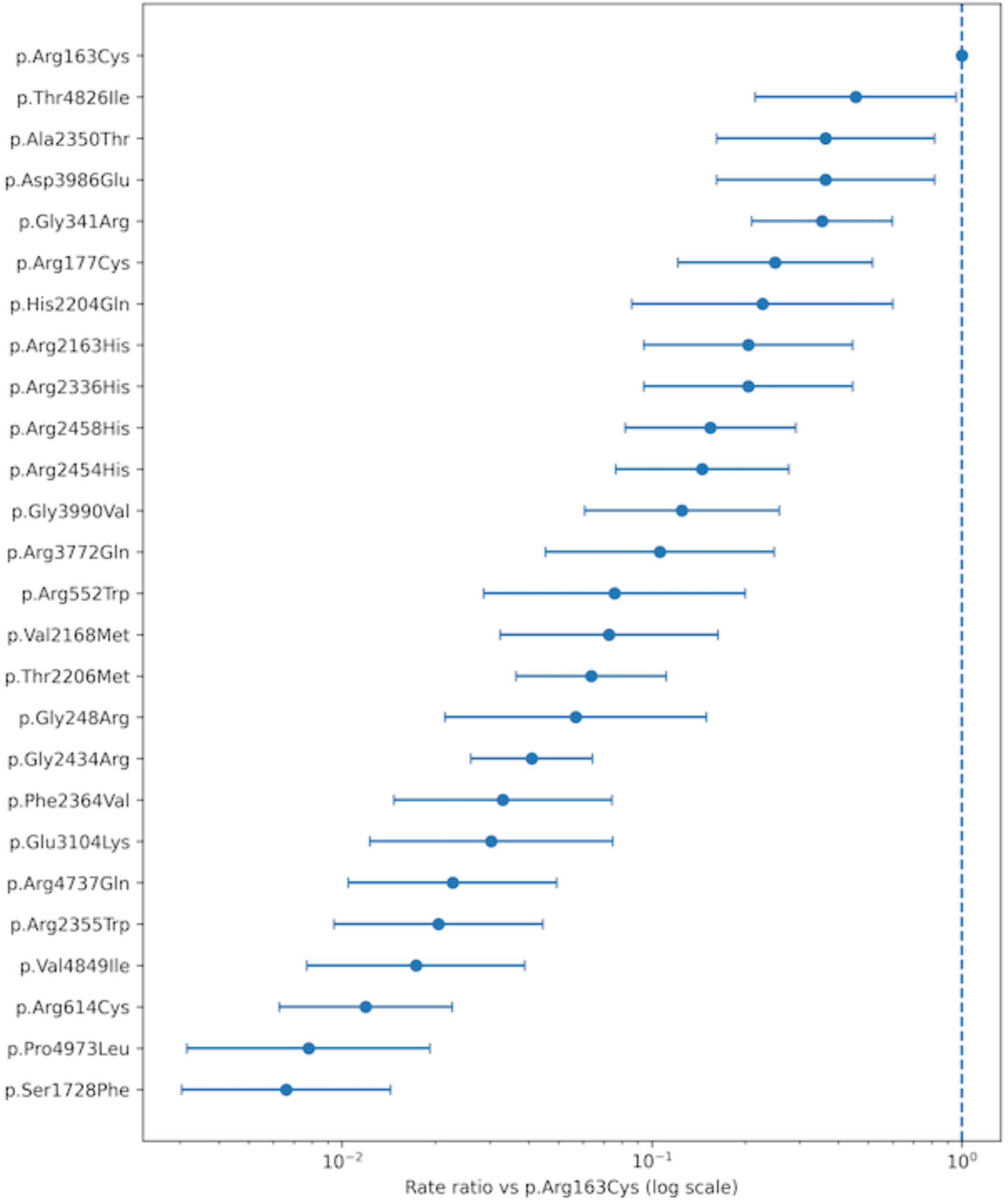
Risk of clinical MH reactions in carriers of 26 *RYR1* variants expressed relative to the risk of carriers of p.Arg163Cys. Values are point estimates and 95% CIs.

### Relative risk of an MH reaction and IVCT responses

The ranked (in descending order) geometric mean responses to the halothane and caffeine contracture tests were obtained for 19 of the *RYR1* variants with relative clinical risk data from Carpenter et al^29^ and they were found to correlate with the relative risk of the variant, (halothane: *ρ* = 0.56, *P* = 0.01 , Fig 3a; caffeine: *ρ* = 0.75, *P* = 0.000145, Fig 3b).

**Figure 3.**
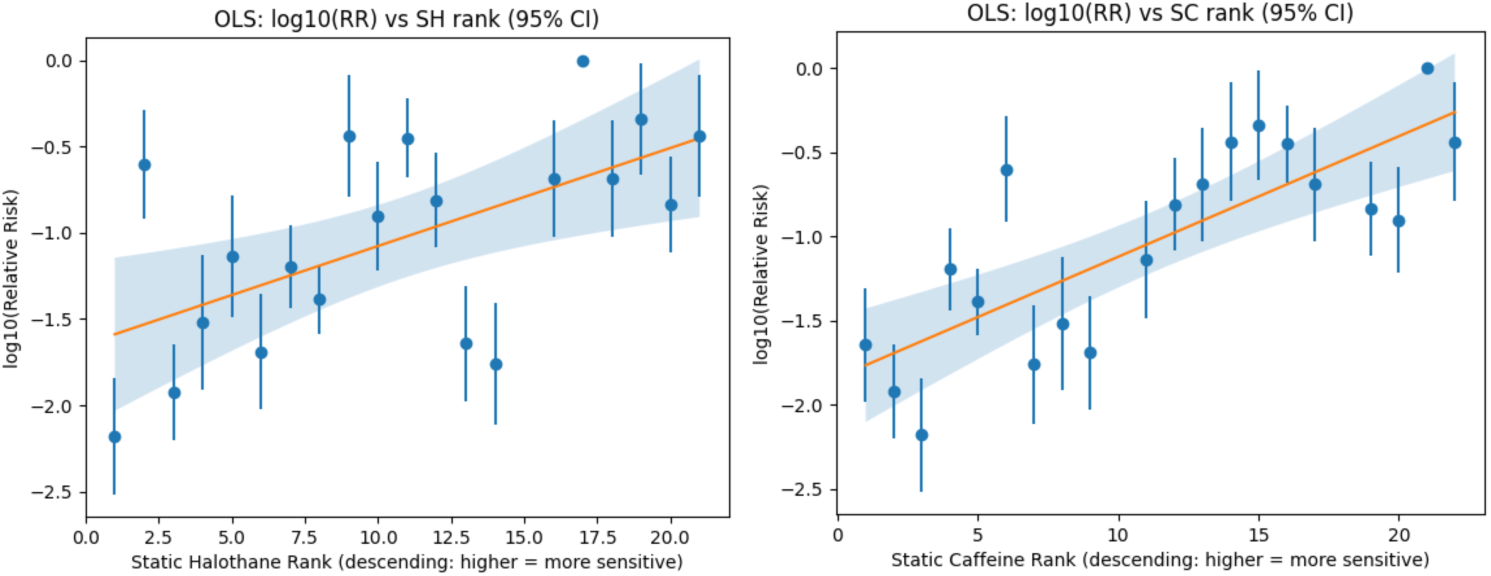
Relationship between relative clinical risk of RYR1 variants and IVCT responses with those variants. The vertical lines indicate 95% confidence interval of relative risk. For illustrative purposes a linear regression line (orange) with 95% confidence interval (shaded area has been fitted). a) static halothane test, b) static caffeine test.

In contrast to the contracture responses, there was a weaker association between the REVEL score (an *in silico* pathogenicity predictor^30^) of each variant and the relative risk (*ρ* = 0.46, *P* = 0.042) but much of the observed association can be attributed to p.Ser1728Phe which has a much lower REVEL score than the other variants (0.477 vs 0.75-0.98). Excluding this variant reduced the degree and significance of the correlation (*ρ* = 0.37, *P* = 0.12).

### *RYR1* genotype and number of general anaesthesia exposures in MH reactors

In addition to examining whether *RYR1* genotype of carriers impacts the risk of developing an MH reaction, we next examined for a relationship between *RYR1* variant and the probability of developing a clinical reaction in those individuals known to have had a reaction (MH reactors). Of the 375 MH index cases with complete anaesthetic history described above, there were 170 who carried 1 of 12 *RYR1* variants found in at least 3 of this cohort. Table 3 shows these variants, the number of reactors with each variant, the number of uneventful exposures in those reactors along with calculated maximum likelihood estimate with 95% credible intervals. We examined the data for evidence that there were differences between the probabilities associated with reactors carrying each variant using binomial general linear modelling. The likelihood ratio of 5.197 (df = 11, *P* = 0.921) indicates that there is no evidence that per- exposure triggering probability is influenced by the RYR1 genotype.

**Table 3.** *RYR1* genotype and anaesthesia exposure history in 170 MH index cases.

| Variant | Uneventful exposures | Events | Total trigger exposures | p_MLE | 95% CrI |
| --- | --- | --- | --- | --- | --- |
| c.7361G>A | 5 | 6 | 11 | 0.55 | 0.28-0.79 |
| c.6617C>T | 13 | 15 | 28 | 0.54 | 0.36-0.71 |
| c.5183C>T | 6 | 6 | 12 | 0.50 | 0.25-0.75 |
| c.1840C>T | 8 | 8 | 16 | 0.50 | 0.28-0.72 |
| c.1021G>A | 16 | 14 | 30 | 0.47 | 0.30-0.64 |
| c.7300G>A | 106 | 81 | 187 | 0.43 | 0.36-0.50 |
| c.11969G>T | 6 | 4 | 10 | 0.40 | 0.17-0.69 |
| c.487C>T | 14 | 9 | 23 | 0.39 | 0.22-0.59 |
| c.529C>T | 10 | 6 | 16 | 0.38 | 0.18-0.62 |
| c.14477C>T | 13 | 7 | 20 | 0.35 | 0.18-0.57 |
| c.7373G>A | 21 | 11 | 32 | 0.34 | 0.20-0.52 |
| c.7304G>A | 7 | 3 | 10 | 0.30 | 0.11-0.61 |

MLE= Maximum likelihood estimation; 95% CrI = 95% credibility interval of the MLE

## Discussion

We have derived a point estimate from clinical epidemiological data for the prevalence of the risk of developing a clinical MH reaction of 1 in 44,000 in the UK population, which is almost 50 times less than the combined prevalence of *RYR1* variants that have been classified as pathogenic or likely pathogenic for MH susceptibility. There have been several previous reports of the prevalence of the risk of developing MH and these have ranged from 1:10,000 – 1:100,000.^31,32^ Previous estimates have been subject to likely errors in the numerator (when suspected cases are not confirmed by IVCT or DNA analysis or not reported), the denominator (uncertainty of general anaesthesia exposure) or both. The foundations of our estimate are robust data derived from a single referral centre operating within a national public health system. These data include the number of confirmed MH events during a period for which we can make an evidence-based informed estimate of the probability of exposure to triggering anaesthetics, along with the risk of an MH reactor reacting with each exposure to general anaesthesia based on the anaesthetic histories of 375 index cases. Interestingly, the derived constant risk of MH reactors developing MH of 2-3% per annum adequately explains the age distribution of MH index cases (most cases occurring before the age of 20, ^33^ at which age the cumulative risk would be 40-60%). Our data do not, however, explain why there are more female than male probands as we found no sex difference in the risk to reactors upon general anaesthesia exposure.^33^

Next, by including event rate and event-reporting data for MH crises over a period of 27 years from a population averaging more than 60 million, we have demonstrated the implausibility of the potential to develop an MH reaction being inherited in an autosomal dominant fashion. This conclusion is based on a conservative estimate of the minimum number of affected individuals (MH reactors) within a family (k=3) for an autosomal dominant trait. It might be argued that k could be 3 or more if the prevalence of the clinical risk was higher and ascertainment (recognition and reporting) of MH cases was lower. However, when we examined this by evaluating k using a prevalence more than double our estimate (1:20,000), a value for k of more than 2, let alone 3 was extremely unlikely. At the same time, this prevalence would imply that the number of true cases of MH 1988-93 was almost 75 with an ascertainment rate of about 33%, which is not supported by anaesthesia mortality data, our national case registry and the fact that only 11 of 789 reactions have occurred in a second individual within a family.

We have previously used genetic data to suggest that MH susceptibility (defined by IVCT phenotype) was unlikely to be uniformly inherited as a monogenic trait.^5^ The current analyses provide independent corroboration in that the collective epidemiology of the more relevant clinical phenotype cannot be explained by an autosomal dominant (monogenic) model. However, the first MH family described by Denborough and colleagues included 10 members who had died under anaesthesia, with an inheritance pattern appropriately described as autosomal dominant with incomplete penetrance (some obligate carriers had received general anaesthesia without reacting).^4^ A situation where inheritance appears monogenic in some families but not in others is best explained by a threshold genetic model. Here, some variants may be so deleterious that they alone are sufficient to disrupt normal function above the threshold whereas other less deleterious variants require to be co-inherited with other functionally relevant variants (in the same or different genes) in order for the threshold to be reached.

We have previously identified differences in IVCT responses and baseline creatine kinase concentration in MH susceptible individuals with different *RYR1* variants.^29^ Using allele frequency data from low-risk UK individuals coupled with UK MH event data we have now demonstrated significant differences in the risk of developing MH among carriers of different *RYR1* variants associated with MH susceptibility, again consistent with a threshold genetic model. Furthermore, the relative risk of a clinical MH reaction correlates with the magnitude of IVCT responses associated with the variant.

It might also have been anticipated that reactors, as well as all carriers, would have a variant-dependent effect on their risk of reacting with each exposure to general anaesthesia but this was not the case. Indeed, any signal that might be gleaned from this small sample shows an inverse relationship. Although the Spearman’s correlation coefficient of -0.58 was non-significant (*P* = 0.062), bootstrap resampling showed that the probability of ρ > 0.2 was < 1%. A similar risk of reactors reacting with each exposure irrespective of the *RYR1* present is consistent with the most straight-forward threshold model where the triggering risk is the same once the combination of contributing genetic factors is sufficiently damaging to reach the threshold (an all-or-none reaction). This is consistent with the overall constant hazard for reacting with each exposure when all 375 reactors were evaluated.

### Implications for family studies, genetic testing and counselling

Following the description of the IVCT, family studies were begun with cascade IVCT testing based on the assumed autosomal dominant inheritance described by Denborough et al.^4^ Early in the history of the IVCT, it appears that the IVCT MHS phenotype was assumed to follow a fully penetrant autosomal dominant inheritance pattern. Reassuringly, later formal evaluation of the IVCT demonstrated its high sensitivity,^9,10^ while early linkage studies used the MHS phenotype to demonstrate linkage initially to chromosome markers and subsequently to specific *RYR1* (and *CACNA1S*) variants,^11–13^ albeit with the observation of frequent examples of genotype-phenotype discordance.^5^ However, while the finding of a pathogenic *RYR1* variant and/or an MHS IVCT phenotype is associated with a risk of developing MH above the background population risk, the current analyses indicate that the elevated risk at the population level may not translate to the individual having a sufficient abnormality for them ever to develop a clinical MH reaction.

Monogenic inheritance is also a basic assumption underlying current variant pathogenicity classification algorithms.^14,15,34^ While pragmatically these classifications may be valid if variation from the expected monogenic inheritance pattern is explained by a “major gene with modifier” scenario, the low relative risk of an MH reaction with some *RYR1* variants suggests that they may not be the largest contributor to the genetic liability to MH within individual patients, i.e., they are unlikely to be the major gene in a “major gene with modifier” scenario. Alternatively, it is perhaps more consistent with our data that they contribute to oligogenic inheritance or even may be a modifier of another, as yet unknown, major gene.

Currently, if there is an incidental finding of an *RYR1* variant classified as pathogenic or likely pathogenic for MH susceptibility, the patient is counselled that they are at risk of developing MH even when there is no suspect anaesthetic history in the family. Furthermore, family cascade testing for the variant is used to label other carriers at increased risk of MH. Our data suggest that their risk is much lower than other risks that are considered to be an indication for IVCT.

While the IVCT remains a highly sensitive test for MH reactors, our analyses suggest that the diagnostic threshold IVCT responses do not adequately discriminate MH reactors from non-reactor *RYR1* variant carriers. In this context, it is worth emphasising that the main clinical value of the IVCT has always been to identify those with an *a priori* increased risk who are actually at no/low risk. Further work is needed to determine if quantitative IVCT responses can be used to distinguish individuals above from those below the clinical reactor threshold.

The incidental finding of *RYR1* missense variants not previously associated with MH also frequently present a clinical conundrum. Often, there is little evidence available to aid variant classification beyond its minor allele frequency in gnomAD and UK Biobank along with its REVEL score: both allele frequency and REVEL score are commonly uninformative. However, the population allele frequencies used to provide evidence against pathogenicity are based on comparison with the allele frequency of known “pathogenic” variants and previously published estimates of the incidence of MH reactions.^14,15^ The current work highlights that while the more prevalent variants associated with MH are important contributors to the population genetic liability to MH, they are likely not major genetic determinants in individual patients because of their low relative risk. More relevant when evaluating a missense *RYR1* variant is how likely is it to be a major genetic determinant for the risk of MH when it has not been previously associated with clinical MH? We can address this question using the results of the simulations presented in Figure 1, from which we can derive mean (95% credibility interval) estimates of the annual risk of an MH reactor having an MH reaction over the past 55 years of 0.021 (0.018-0.024). If we consider a minimum “penetrance” of 0.01 for a variant to be considered a major genetic determinant, we can calculate a threshold allele prevalence of 2.6 x 10^-6^ (2.3 - 3.0 x 10^-6^) above which a variant would be expected to have been associated with one or more MH reactions over the past 55 years with ≥ 97.5% (two-tailed) probability. We therefore recommend that variants with a minor allele frequency > 3.0 x 10^-6^ in UK Biobank (an allele count of > 2) that have not been associated with a MH reaction in the UK can be excluded from being a major genetic determinant of MH.

### Limitations

These analyses are based on the assumptions that the proportion of MH reactors in the UK population has remained constant over the past 55 years and all members of the population (including all carriers & reactors) have an equal chance of exposure to anaesthetics until they are flagged as potentially susceptible to MH through a personal or family history of a suspected reaction. While there is no reason to suspect that the latter assumption is invalid, the proportion of MH reactors might have changed as a result of the considerable net migration into the UK over the 55 year period.^26^ However, we consider that any such change would lie within the tolerance limits of our statistical models. We have also assumed that the ethnicity profiles of our cohort and of UK Biobank participants match the UK population. The non-white proportion of our cohort is 5%, which is similar to the 5-6% proportion in UK Biobank^35^. The latest estimates for the UK population (2021 census^36^) however have a higher non-white proportion of 18%. These disparities probably arise from the dates of sampling of the UK MH (1971-25, 86% of index case reactions occurred before 2011) and UK Biobank (2006-10) cohorts, reflecting less population diversity prior to 2011 compared with 2021. These data indicate that the UK Biobank resource is a very reasonable low-risk comparator for derivation of *RYR1* variant carrier prevalence to be used with MH cohort genotype data. Matching of the ethnicity (albeit crudely as white and non-white groups) between the MH and Biobank cohorts also provides no evidence that the prevalence of the risk of MH varies between ethnic groups. It is likely, however, that the contributions of specific *RYR1* variants to the overall risk will be ethnicity-dependent.^37–42^

## Conclusion

The prevalence of those at risk of developing MH in the UK is lower than the consensus estimate of 1:10,000^31^ and considerably lower than the combined prevalence of *RYR1* variants classified as pathogenic or likely pathogenic (1:900). The contribution to the clinical risk of MH varies markedly between these variants. These findings add significant weight in support of a threshold inheritance model. They also suggest that current diagnostics, both genotyping and IVCT phenotyping, over-diagnose the clinical risk. Genome-wide analyses of large cohorts of MH reactors, variant carriers, MH (IVCT) negative relatives and low-risk controls are needed to identify additional genes and variants contributing to the genetic liability of MH.

## Data Availability

All data produced in the present study and not contained in the manuscript or supplementary files are available upon reasonable request to the authors

## Author Contributions

Study concept and design: PMH, EMA

Data acquisition: EMA, CD, NF, SH, HG, JBG, PKG, PMH, SS, RLR

Data analysis: PMH, EMA

Data interpretation: All authors

Drafting manuscript: PMH

Critical review of manuscript: EMA, CD, NF, SH, HG, JBG, PKG, M-AS, SS, RLR

Approval of final draft: All authors

## Funding

Genetic data included in this study have been collected over the past 35 years. The following grants have contributed to the funding of this work:

UK Department of Health Pharmacogenetics Research Grant Programme. (PMH, MAS, RLR). 2004-2008

Big Lottery Fund (RLR, PMH). 2005-2008

National Institutes of Health; US National Institute of Arthritis and Musculoskeletal and Skin Diseases (2P01 AR-05235 to PMH). 2012-2017

The opinions expressed in this article are the authors’ own and do not reflect the view of the National Institutes of Health, the Department of Health and Human Services or the United States government. The content is solely the responsibility of the authors and does not necessarily represent the official views of the National Institutes of Health.

## List of Supplementary Tables

Supplementary table S1. The number of MH reactions in the UK and number of referrals to the UK Malignant Hyperthermia Unit 1988-2025

Supplementary table S2. Proportion of male and female MH index cases reacting with each general anaesthesia exposure

Supplementary table S3. RYR1 variant allele frequencies in the UK Biobank whole genome sequence cohort and the non-UK Biobank subset of the non-Finnish European cohort of gnomAd (version 4.1)

Supplementary Table S4 (separate PDF file). Pairwise comparison of the relative risk of carriers of 26 RYR1 variants developing clinical malignant hyperthermia (MH).

## References

1. Rüffert H, Gillies R, Hopkins PM, et al. European Malignant Hyperthermia Group 2025 guidelines for the investigation of malignant hyperthermia susceptibility. Br J Anaesth. 2026;136:653–661. PMID: 41478797

2. Hopkins PM, Girard T, Dalay S, Jenkins B, Thacker A, Patteril M, McGrady E. Malignant hyperthermia 2020: Guideline from the Association of Anaesthetists. Anaesthesia. 2021;76:655–64

3. Glahn KPE, Girard T, Hellblom A, et al. Recognition and management of a malignant hyperthermia crisis: updated 2024 guideline from the European Malignant Hyperthermia Group. Br J Anaesth. 2025;134:221–223. PMID: 39482150

4. Denborough MA, Forster JF, Lovell RR, Maplestone PA, Villiers JD. Anaesthetic deaths in a family. Br J Anaesth. 1962;34:395–6. PMID: 13885389.

5. Miller DM, Daly C, Aboelsaod EM, et al. Genetic epidemiology of malignant hyperthermia in the UK. Br J Anaesth. 2018;121:944–952. PMID: 30236257

6. Hopkins PM. What is malignant hyperthermia susceptibility? Br J Anaesth. 2023;131:5–8

7. Kalow W, Britt BA, Terreau ME, Haist C. Metabolic error of muscle metabolism after recovery from malignant hyperthermia. Lancet. 1970;2(7679):895-. PMID: 4097281.

8. Ellis FR, Harriman DG, Keaney NP, Kyei-Mensah K, Tyrrell JH. Halothane-induced muscle contracture as a cause of hyperpyrexia. Br J Anaesth. 1971;43:721–2. PMID: 5564247.

9. Ording H, Brancadoro V, Cozzolino S, et al. In vitro contracture test for diagnosis of malignant hyperthermia following the protocol of the European MH Group: results of testing patients surviving fulminant MH and unrelated low-risk subjects. Acta Anaesthesiol Scand. 1997;41:955–66. PMID: 9311391.

10. Hopkins PM, Rüffert H, Snoeck MM, et al. European Malignant Hyperthermia Group guidelines for investigation of malignant hyperthermia susceptibility. Br J Anaesth. 2015;115:531–9. PMID: 26188342.

11. MacLennan DH, Duff C, Zorzato F, et al. Ryanodine receptor gene is a candidate for predisposition to malignant hyperthermia. Nature. 1990;343:559–61. PMID: 1967823.

12. McCarthy TV, Healy JM, Heffron JJ, et al. Localization of the malignant hyperthermia susceptibility locus to human chromosome 19q12-13.2. Nature. 1990;343:562–4. PMID: 2300206.

13. Robinson RL, Monnier N, Wolz W, et al. A genome wide search for susceptibility loci in three European malignant hyperthermia pedigrees. Hum Mol Genet. 1997;6:953–61. PMID: 9175745.

14. Johnston JJ, Dirksen RT, Girard T, et al. Variant curation expert panel recommendations for RYR1 pathogenicity classifications in malignant hyperthermia susceptibility. Genet Med. 2021;23:1288–1295. PMID: 33767344

15. Johnston JJ, Dirksen RT, Girard T, et al. Updated variant curation expert panel criteria and pathogenicity classifications for 251 variants for RYR1-related malignant hyperthermia susceptibility. Hum Mol Genet. 2022;31:4087–4093. PMID: 35849058.

16. Deufel, T, Sudbrak, R, Feist, Y, Rubsam, B, Du Chesne, I, Schafer, KL et al. Discordance, in a malignant hyperthermia pedigree, between in vitro contracture-test phenotypes and haplotypes for the MHS1 region on chromosome 19q12-13.2, comprising the C1840T transition in the RYR1 gene. Am J Hum Genet. 1995; 56:1334–1342.

17. Fagerlund, TH, Ording, H, Bendixen, D, Islander, G, Ranklev Twetman, E, Berg, K. Discordance between malignant hyperthermia susceptibility and RYR1 mutation C1840T in two Scandinavian MH families exhibiting this mutation. Clin Genet. 1997; 52:416–421.

18. Adeokun AM, West SP, Ellis FR, et al. The G1021A substitution in the RYR1 gene does not always cosegregate with malignant hyperthermia susceptibility in a British pedigree. American Journal of Human Genetics 1997; 60: 833–841.

19. Robinson RL, Anetseder MJ, Brancadoro V, et al. Recent advances in the diagnosis of malignant hyperthermia susceptibility: how confident can we be of genetic testing? European Journal of Human Genetics 2003;11:342–8

20. Campling EA, Devlin HB, Hoile RW, Lunn JN. The Report of the National Confidential Enquiry into Perioperative Deaths 1990. https://www.ncepod.org.uk/1990report/Full%20Report%201990.pdf (accessed 19/11/25)

21. Office for National Statistics. Birth summary tables: Live births in England and Wales. https://www.ons.gov.uk/peoplepopulationandcommunity/birthsdeathsandmarriages/livebirths/datasets/birthsummarytables (accessed 20/11/25)

22. Francome C, Savage W. Caesarean section in Britain and the United States: 12% or 24%? Soc Sci Med. 1993;37:1199–218.

23. UK Parliament. Caesarean births: Written answers, House of Lords (HL Deb 19 July 1999). https://api.parliament.uk/historic-hansard/written-answers/1999/jul/19/caesarean-births (accessed 20/11/25)

24. Nicholl JP, Williams BT, Beeby NR. Role of the private sector in elective surgery in England and Wales, 1986. BMJ. 1989;298:243–7

25. Sury MRJ, Palmer JHMG, Cook TM, Pandit JJ, Mahajan RP. The state of UK anaesthesia: A survey of National Health Service activity in 2013. Br J Anaesth. 2014;113:575–84

26. Office for National Statistics. UK population mid-year estimate time series. https://www.ons.gov.uk/peoplepopulationandcommunity/populationandmigration/populationestimates/timeseries/ukpop/pop (accessed 20/11/25)

27. Woodall NM, Cook TM; National Audit Project 4 investigators. National census of airway management techniques used for anaesthesia in the UK: First phase of the 4th National Audit Project. Br J Anaesth. 2011;106(2):266–71.

28. Kemp H, Marinho S, Cook TM, et al. An observational national study of anaesthetic workload and seniority across the working week and weekend in the UK in 2016: the 6th National Audit Project (NAP6) Activity Survey. Br J Anaesth. 2018;121:134–145. PMID: 29935565.

29. Carpenter D, Robinson RL, Quinnell RJ, Ringrose C, Hogg M, Casson F, Booms P, Iles D, Halsall PJ, Steele D, Shaw M-A, Hopkins PM. Genetic variation in RYR1 and malignant hyperthermia phenotypes. British Journal of Anaesthesia 2009; 103: 538–548

30. Ioannidis NM, Rothstein JH, Pejaver V, et al. REVEL: An Ensemble Method for Predicting the Pathogenicity of Rare Missense Variants. Am J Hum Genet. 2016;99:877–885. PMID: 27666373

31. Hopkins PM. Malignant hyperthermia. Curr Anaesth Crit Care 2008; 19: 22–33

32. Brady JE, Sun LS, Rosenberg H, Li G. Prevalence of malignant hyperthermia due to anesthesia in New York State, 2001-2005. Anesth Analg 2009; 109: 1162–6

33. Gupta PK, Hopkins PM. Diagnosis and management of malignant hyperthermia. BJA Education 2017; 17: 249–54

34. Richards S, Aziz N, Bale S, et al. Standards and guidelines for the interpretation of sequence variants: a joint consensus recommendation of the American College of Medical Genetics and Genomics and the Association for Molecular Pathology. Genet Med. 2015;17:405–24. PMID: 25741868

35. Bycroft, C., Freeman, C., Petkova, D. et al. The UK Biobank resource with deep phenotyping and genomic data. Nature 2018:562; 203–209

36. Office for National Statistics. Ethnic Group, England and Wales: Census 2021. https://www.ons.gov.uk/peoplepopulationandcommunity/culturalidentity/ethnicity/bulletins/ethnicgroupenglandandwales/census2021#

37. Ibarra M CA, Wu S, Murayama K, Minami N, Ichihara Y, Kikuchi H, Noguchi S, Hayashi YK, Ochiai R, Nishino I. Malignant hyperthermia in Japan: mutation screening of the entire ryanodine receptor type 1 gene coding region by direct sequencing. Anesthesiology 2006;104:1146–54.

38. Galli L, Orrico A, Lorenzini S, et al. Frequency and localization of mutations in the 106 exons of the RYR1 gene in 50 individuals with malignant hyperthermia. Hum Mutat 2006;27:830

39. Broman M, Gehrig A, Islander G, et al. Mutation screening of the RYR1-cDNA from peripheral B-lymphocytes in 15 Swedish malignant hyperthermia index cases. Br J Anaesth 2009;102:642–9.

40. Kraeva N, Riazi, S, Loke J, et al. Ryanodine receptor type 1 gene mutations found in the Canadian malignant hyperthermia population. Can J Anesth 2011; 58:504–13

41. Brandom BW, Bina S, Wong CA, et al. Ryanodine receptor type 1 gene variants in the malignant hyperthermia-susceptible population of the United States. Anesth Analg 2013; 116: 1078–86

42. Gillies RL, Bjorksten AR, Du Sart D, Hockey BM. Analysis of the entire ryanodine receptor type 1 and alpha 1 subunit of the dihydropyridine receptor (CACNA1S) coding regions for variants associated with malignant hyperthermia in Australian families. Anaesth Intensive Care 2015; 43: 157–66

