## Supplementary Table S4 for "Epidemiology of malignant hyperthermia in the UK 1988-2025: implications for prevalence, mode of inheritance, relative risk associated with *RYR1* genotypes and *in vitro* contracture test phenotype"

| | A | B | C | D | E | F | G | H | I | J | K | L |
|---|---|---|---|---|---|---|---|---|---|---|---|---|
| 1 | **variant_A** | **variant_B** | **events_A** | **carrier_yrs_A** | **rate_A** | **events_B** | **carrier_yrs_B** | **rate_B** | **RR_A_over_B** | **LR_statistic** | **p_value_LR** | **p_adj_FDR_BH** |
| 2 | p.Arg163Cys | p.Gly341Arg | 22 | 6953 | 0.00316 | 39 | 34757 | 0.00112 | 2.82 | 13.29 | 0.000266878 | 0.000682956 |
| 3 | p.Arg163Cys | p.Ala2350Thr | 22 | 6953 | 0.00316 | 8 | 6951 | 0.00115 | 2.75 | 6.79 | 0.009164592 | 0.016455758 |
| 4 | p.Arg163Cys | p.Thr4826Ile | 22 | 6953 | 0.00316 | 10 | 6951 | 0.00144 | 2.20 | 4.61 | 0.031803573 | 0.05091705 |
| 5 | p.Arg163Cys | p.Arg2163His | 22 | 6953 | 0.00316 | 9 | 13903 | 0.00065 | 4.89 | 18.28 | 1.91E-05 | 5.85E-05 |
| 6 | p.Arg163Cys | p.Arg2336His | 22 | 6953 | 0.00316 | 9 | 13903 | 0.00065 | 4.89 | 18.28 | 1.91E-05 | 5.85E-05 |
| 7 | p.Arg163Cys | p.Arg177Cys | 22 | 6953 | 0.00316 | 11 | 13904 | 0.00079 | 4.00 | 15.25 | 9.44E-05 | 0.000266715 |
| 8 | p.Arg163Cys | p.Arg2458His | 22 | 6953 | 0.00316 | 17 | 34758 | 0.00049 | 6.47 | 31.61 | 1.89E-08 | 1.12E-07 |
| 9 | p.Arg163Cys | p.Arg2454His | 22 | 6953 | 0.00316 | 16 | 34757 | 0.00046 | 6.87 | 32.94 | 9.52E-09 | 5.73E-08 |
| 10 | p.Arg163Cys | p.Gly3990Val | 22 | 6953 | 0.00316 | 11 | 27806 | 0.00040 | 8.00 | 33.71 | 6.41E-09 | 4.25E-08 |
| 11 | p.Arg163Cys | p.Val2168Met | 22 | 6953 | 0.00316 | 8 | 34757 | 0.00023 | 13.75 | 46.95 | 7.28E-12 | 8.15E-11 |
| 12 | p.Arg163Cys | p.Gly248Arg | 22 | 6953 | 0.00316 | 5 | 27807 | 0.00018 | 17.60 | 47.17 | 6.52E-12 | 7.85E-11 |
| 13 | p.Arg163Cys | p.Thr2206Met | 22 | 6953 | 0.00316 | 28 | 139031 | 0.00020 | 15.71 | 68.09 | 1.56E-16 | 2.99E-15 |
| 14 | p.Arg163Cys | p.Gly2434Arg | 22 | 6953 | 0.00316 | 127 | 980149 | 0.00013 | 24.42 | 95.09 | 1.82E-22 | 6.55E-21 |
| 15 | p.Arg163Cys | p.Glu3104Lys | 22 | 6953 | 0.00316 | 6 | 62563 | 0.00010 | 32.99 | 73.47 | 1.02E-17 | 2.07E-16 |
| 16 | p.Arg163Cys | p.Arg4737Gln | 22 | 6953 | 0.00316 | 9 | 125128 | 0.00007 | 43.99 | 93.17 | 4.80E-22 | 1.56E-20 |
| 17 | p.Arg163Cys | p.Arg2355Trp | 22 | 6953 | 0.00316 | 9 | 139029 | 0.00006 | 48.88 | 97.48 | 5.45E-23 | 2.21E-21 |
| 18 | p.Arg163Cys | p.Val4849Ile | 22 | 6953 | 0.00316 | 8 | 145983 | 0.00005 | 57.74 | 101.95 | 5.71E-24 | 2.65E-22 |
| 19 | p.Arg163Cys | p.Ser1728Phe | 22 | 6953 | 0.00316 | 9 | 430988 | 0.00002 | 151.52 | 145.22 | 1.92E-33 | 3.12E-31 |
| 20 | p.Arg163Cys | p.Arg614Cys | 22 | 6953 | 0.00316 | 16 | 424097 | 0.00004 | 83.87 | 130.38 | 3.38E-30 | 2.75E-28 |
| 21 | p.Arg163Cys | p.Asp3986Glu | 22 | 6953 | 0.00316 | 8 | 6951 | 0.00115 | 2.75 | 6.79 | 0.009164006 | 0.016455758 |
| 22 | p.Arg163Cys | p.His2204Gln | 22 | 6953 | 0.00316 | 5 | 6952 | 0.00072 | 4.40 | 11.55 | 0.000677008 | 0.001582932 |
| 23 | p.Arg163Cys | p.Arg552Trp | 22 | 6953 | 0.00316 | 5 | 20856 | 0.00024 | 13.20 | 37.99 | 7.10E-10 | 5.36E-09 |
| 24 | p.Arg163Cys | p.Arg3772Gln | 22 | 6953 | 0.00316 | 7 | 20855 | 0.00034 | 9.43 | 32.96 | 9.39E-09 | 5.73E-08 |
| 25 | p.Arg163Cys | p.Phe2364Val | 22 | 6953 | 0.00316 | 8 | 76478 | 0.00010 | 30.25 | 75.93 | 2.94E-18 | 6.37E-17 |
| 26 | p.Arg163Cys | p.Pro4973Leu | 22 | 6953 | 0.00316 | 6 | 243302 | 0.00002 | 128.30 | 128.91 | 7.11E-30 | 4.62E-28 |
| 27 | p.Gly341Arg | p.Ala2350Thr | 39 | 34757 | 0.00112 | 8 | 6951 | 0.00115 | 0.98 | 0.00 | 0.948142879 | 0.95403047 |
| 28 | p.Gly341Arg | p.Thr4826Ile | 39 | 34757 | 0.00112 | 10 | 6951 | 0.00144 | 0.78 | 0.47 | 0.494213659 | 0.546324623 |
| 29 | p.Gly341Arg | p.Arg2163His | 39 | 34757 | 0.00112 | 9 | 13903 | 0.00065 | 1.73 | 2.47 | 0.116249215 | 0.158743676 |
| 30 | p.Gly341Arg | p.Arg2336His | 39 | 34757 | 0.00112 | 9 | 13903 | 0.00065 | 1.73 | 2.47 | 0.116237841 | 0.158743676 |
| 31 | p.Gly341Arg | p.Arg177Cys | 39 | 34757 | 0.00112 | 11 | 13904 | 0.00079 | 1.42 | 1.12 | 0.290887982 | 0.350142941 |
| 32 | p.Gly341Arg | p.Arg2458His | 39 | 34757 | 0.00112 | 17 | 34758 | 0.00049 | 2.29 | 8.88 | 0.002882301 | 0.005891496 |
| 33 | p.Gly341Arg | p.Arg2454His | 39 | 34757 | 0.00112 | 16 | 34757 | 0.00046 | 2.44 | 9.92 | 0.001634466 | 0.003565111 |

| | A | B | C | D | E | F | G | H | I | J | K | L |
|---|---|---|---|---|---|---|---|---|---|---|---|---|
| 34 | p.Gly341Arg | p.Gly3990Val | 39 | 34757 | 0.00112 | 11 | 27806 | 0.00040 | 2.84 | 11.00 | 0.000912468 | 0.002073791 |
| 35 | p.Gly341Arg | p.Val2168Met | 39 | 34757 | 0.00112 | 8 | 34757 | 0.00023 | 4.88 | 22.27 | 2.37E-06 | 9.27E-06 |
| 36 | p.Gly341Arg | p.Gly248Arg | 39 | 34757 | 0.00112 | 5 | 27807 | 0.00018 | 6.24 | 22.80 | 1.80E-06 | 7.21E-06 |
| 37 | p.Gly341Arg | p.Thr2206Met | 39 | 34757 | 0.00112 | 28 | 139031 | 0.00020 | 5.57 | 46.97 | 7.22E-12 | 8.15E-11 |
| 38 | p.Gly341Arg | p.Gly2434Arg | 39 | 34757 | 0.00112 | 127 | 980149 | 0.00013 | 8.66 | 91.04 | 1.41E-21 | 4.16E-20 |
| 39 | p.Gly341Arg | p.Glu3104Lys | 39 | 34757 | 0.00112 | 6 | 62563 | 0.00010 | 11.70 | 50.27 | 1.34E-12 | 1.89E-11 |
| 40 | p.Gly341Arg | p.Arg4737Gln | 39 | 34757 | 0.00112 | 9 | 125128 | 0.00007 | 15.60 | 77.12 | 1.61E-18 | 3.74E-17 |
| 41 | p.Gly341Arg | p.Arg2355Trp | 39 | 34757 | 0.00112 | 9 | 139029 | 0.00006 | 17.33 | 83.23 | 7.32E-20 | 1.83E-18 |
| 42 | p.Gly341Arg | p.Val4849Ile | 39 | 34757 | 0.00112 | 8 | 145983 | 0.00005 | 20.48 | 89.13 | 3.70E-21 | 1.00E-19 |
| 43 | p.Gly341Arg | p.Ser1728Phe | 39 | 34757 | 0.00112 | 9 | 430988 | 0.00002 | 53.73 | 157.50 | 3.98E-36 | 1.29E-33 |
| 44 | p.Gly341Arg | p.Arg614Cys | 39 | 34757 | 0.00112 | 16 | 424097 | 0.00004 | 29.74 | 137.46 | 9.55E-32 | 1.04E-29 |
| 45 | p.Gly341Arg | p.Asp3986Glu | 39 | 34757 | 0.00112 | 8 | 6951 | 0.00115 | 0.98 | 0.00 | 0.948159513 | 0.95403047 |
| 46 | p.Gly341Arg | p.His2204Gln | 39 | 34757 | 0.00112 | 5 | 6952 | 0.00072 | 1.56 | 0.98 | 0.321622373 | 0.382883777 |
| 47 | p.Gly341Arg | p.Arg552Trp | 39 | 34757 | 0.00112 | 5 | 20856 | 0.00024 | 4.68 | 15.31 | 9.10E-05 | 0.00025951 |
| 48 | p.Gly341Arg | p.Arg3772Gln | 39 | 34757 | 0.00112 | 7 | 20855 | 0.00034 | 3.34 | 11.16 | 0.000836692 | 0.001914964 |
| 49 | p.Gly341Arg | p.Phe2364Val | 39 | 34757 | 0.00112 | 8 | 76478 | 0.00010 | 10.73 | 53.84 | 2.17E-13 | 3.53E-12 |
| 50 | p.Gly341Arg | p.Pro4973Leu | 39 | 34757 | 0.00112 | 6 | 243302 | 0.00002 | 45.50 | 128.46 | 8.91E-30 | 4.82E-28 |
| 51 | p.Ala2350Thr | p.Thr4826Ile | 8 | 6951 | 0.00115 | 10 | 6951 | 0.00144 | 0.80 | 0.22 | 0.637019537 | 0.68327178 |
| 52 | p.Ala2350Thr | p.Arg2163His | 8 | 6951 | 0.00115 | 9 | 13903 | 0.00065 | 1.78 | 1.37 | 0.242153672 | 0.298113909 |
| 53 | p.Ala2350Thr | p.Arg2336His | 8 | 6951 | 0.00115 | 9 | 13903 | 0.00065 | 1.78 | 1.37 | 0.242140574 | 0.298113909 |
| 54 | p.Ala2350Thr | p.Arg177Cys | 8 | 6951 | 0.00115 | 11 | 13904 | 0.00079 | 1.45 | 0.63 | 0.425730816 | 0.483796286 |
| 55 | p.Ala2350Thr | p.Arg2458His | 8 | 6951 | 0.00115 | 17 | 34758 | 0.00049 | 2.35 | 3.52 | 0.060497056 | 0.090193967 |
| 56 | p.Ala2350Thr | p.Arg2454His | 8 | 6951 | 0.00115 | 16 | 34757 | 0.00046 | 2.50 | 3.95 | 0.046875747 | 0.0715268 |
| 57 | p.Ala2350Thr | p.Gly3990Val | 8 | 6951 | 0.00115 | 11 | 27806 | 0.00040 | 2.91 | 4.80 | 0.028520999 | 0.046348466 |
| 58 | p.Ala2350Thr | p.Val2168Met | 8 | 6951 | 0.00115 | 8 | 34757 | 0.00023 | 5.00 | 9.40 | 0.002164356 | 0.004627955 |
| 59 | p.Ala2350Thr | p.Gly248Arg | 8 | 6951 | 0.00115 | 5 | 27807 | 0.00018 | 6.40 | 10.66 | 0.001094881 | 0.002437351 |
| 60 | p.Ala2350Thr | p.Thr2206Met | 8 | 6951 | 0.00115 | 28 | 139031 | 0.00020 | 5.71 | 13.31 | 0.000264559 | 0.000682432 |
| 61 | p.Ala2350Thr | p.Gly2434Arg | 8 | 6951 | 0.00115 | 127 | 980149 | 0.00013 | 8.88 | 20.36 | 6.42E-06 | 2.29E-05 |
| 62 | p.Ala2350Thr | p.Glu3104Lys | 8 | 6951 | 0.00115 | 6 | 62563 | 0.00010 | 12.00 | 18.98 | 1.32E-05 | 4.31E-05 |
| 63 | p.Ala2350Thr | p.Arg4737Gln | 8 | 6951 | 0.00115 | 9 | 125128 | 0.00007 | 16.00 | 24.58 | 7.14E-07 | 2.98E-06 |
| 64 | p.Ala2350Thr | p.Arg2355Trp | 8 | 6951 | 0.00115 | 9 | 139029 | 0.00006 | 17.78 | 26.08 | 3.27E-07 | 1.46E-06 |
| 65 | p.Ala2350Thr | p.Val4849Ile | 8 | 6951 | 0.00115 | 8 | 145983 | 0.00005 | 21.00 | 28.02 | 1.20E-07 | 5.91E-07 |
| 66 | p.Ala2350Thr | p.Ser1728Phe | 8 | 6951 | 0.00115 | 9 | 430988 | 0.00002 | 55.11 | 43.07 | 5.28E-11 | 5.20E-10 |

|  | A | B | C | D | E | F | G | H | I | J | K | L |
|---|---|---|---|---|---|---|---|---|---|---|---|---|
| 67 | p.Ala2350Thr | p.Arg614Cys | 8 | 6951 | 0.00115 | 16 | 424097 | 0.00004 | 30.50 | 36.00 | 1.97E-09 | 1.39E-08 |
| 68 | p.Ala2350Thr | p.Asp3986Glu | 8 | 6951 | 0.00115 | 8 | 6951 | 0.00115 | 1.00 | 0.00 | 0.999986988 | 0.999986988 |
| 69 | p.Ala2350Thr | p.His2204Gln | 8 | 6951 | 0.00115 | 5 | 6952 | 0.00072 | 1.60 | 0.70 | 0.403227949 | 0.464722551 |
| 70 | p.Ala2350Thr | p.Arg552Trp | 8 | 6951 | 0.00115 | 5 | 20856 | 0.00024 | 4.80 | 7.74 | 0.005415627 | 0.010415114 |
| 71 | p.Ala2350Thr | p.Arg3772Gln | 8 | 6951 | 0.00115 | 7 | 20855 | 0.00034 | 3.43 | 5.48 | 0.019226816 | 0.03278683 |
| 72 | p.Ala2350Thr | p.Phe2364Val | 8 | 6951 | 0.00115 | 8 | 76478 | 0.00010 | 11.00 | 18.97 | 1.33E-05 | 4.31E-05 |
| 73 | p.Ala2350Thr | p.Pro4973Leu | 8 | 6951 | 0.00115 | 6 | 243302 | 0.00002 | 46.67 | 38.55 | 5.33E-10 | 4.33E-09 |
| 74 | p.Thr4826Ile | p.Arg2163His | 10 | 6951 | 0.00144 | 9 | 13903 | 0.00065 | 2.22 | 2.98 | 0.08411246 | 0.11843998 |
| 75 | p.Thr4826Ile | p.Arg2336His | 10 | 6951 | 0.00144 | 9 | 13903 | 0.00065 | 2.22 | 2.98 | 0.084106246 | 0.11843998 |
| 76 | p.Thr4826Ile | p.Arg177Cys | 10 | 6951 | 0.00144 | 11 | 13904 | 0.00079 | 1.82 | 1.83 | 0.176308147 | 0.229200591 |
| 77 | p.Thr4826Ile | p.Arg2458His | 10 | 6951 | 0.00144 | 17 | 34758 | 0.00049 | 2.94 | 6.44 | 0.011158433 | 0.019816889 |
| 78 | p.Thr4826Ile | p.Arg2454His | 10 | 6951 | 0.00144 | 16 | 34757 | 0.00046 | 3.13 | 7.02 | 0.008046929 | 0.014692426 |
| 79 | p.Thr4826Ile | p.Gly3990Val | 10 | 6951 | 0.00144 | 11 | 27806 | 0.00040 | 3.64 | 8.03 | 0.004592447 | 0.008937397 |
| 80 | p.Thr4826Ile | p.Val2168Met | 10 | 6951 | 0.00144 | 8 | 34757 | 0.00023 | 6.25 | 14.02 | 0.000180717 | 0.000485396 |
| 81 | p.Thr4826Ile | p.Gly248Arg | 10 | 6951 | 0.00144 | 5 | 27807 | 0.00018 | 8.00 | 15.33 | 9.05E-05 | 0.00025951 |
| 82 | p.Thr4826Ile | p.Thr2206Met | 10 | 6951 | 0.00144 | 28 | 139031 | 0.00020 | 7.14 | 19.82 | 8.50E-06 | 2.94E-05 |
| 83 | p.Thr4826Ile | p.Gly2434Arg | 10 | 6951 | 0.00144 | 127 | 980149 | 0.00013 | 11.10 | 29.31 | 6.16E-08 | 3.18E-07 |
| 84 | p.Thr4826Ile | p.Glu3104Lys | 10 | 6951 | 0.00144 | 6 | 62563 | 0.00010 | 15.00 | 26.15 | 3.17E-07 | 1.45E-06 |
| 85 | p.Thr4826Ile | p.Arg4737Gln | 10 | 6951 | 0.00144 | 9 | 125128 | 0.00007 | 20.00 | 33.58 | 6.86E-09 | 4.46E-08 |
| 86 | p.Thr4826Ile | p.Arg2355Trp | 10 | 6951 | 0.00144 | 9 | 139029 | 0.00006 | 22.22 | 35.48 | 2.57E-09 | 1.78E-08 |
| 87 | p.Thr4826Ile | p.Val4849Ile | 10 | 6951 | 0.00144 | 8 | 145983 | 0.00005 | 26.25 | 37.83 | 7.70E-10 | 5.69E-09 |
| 88 | p.Thr4826Ile | p.Ser1728Phe | 10 | 6951 | 0.00144 | 9 | 430988 | 0.00002 | 68.89 | 56.86 | 4.67E-14 | 8.43E-13 |
| 89 | p.Thr4826Ile | p.Arg614Cys | 10 | 6951 | 0.00144 | 16 | 424097 | 0.00004 | 38.13 | 48.42 | 3.44E-12 | 4.47E-11 |
| 90 | p.Thr4826Ile | p.Asp3986Glu | 10 | 6951 | 0.00144 | 8 | 6951 | 0.00115 | 1.25 | 0.22 | 0.637007202 | 0.68327178 |
| 91 | p.Thr4826Ile | p.His2204Gln | 10 | 6951 | 0.00144 | 5 | 6952 | 0.00072 | 2.00 | 1.70 | 0.192406727 | 0.24618971 |
| 92 | p.Thr4826Ile | p.Arg552Trp | 10 | 6951 | 0.00144 | 5 | 20856 | 0.00024 | 6.00 | 11.51 | 0.00069287 | 0.001608448 |
| 93 | p.Thr4826Ile | p.Arg3772Gln | 10 | 6951 | 0.00144 | 7 | 20855 | 0.00034 | 4.29 | 8.72 | 0.003149469 | 0.006357624 |
| 94 | p.Thr4826Ile | p.Phe2364Val | 10 | 6951 | 0.00144 | 8 | 76478 | 0.00010 | 13.75 | 26.36 | 2.83E-07 | 1.31E-06 |
| 95 | p.Thr4826Ile | p.Pro4973Leu | 10 | 6951 | 0.00144 | 6 | 243302 | 0.00002 | 58.33 | 50.84 | 1.00E-12 | 1.48E-11 |
| 96 | p.Arg2163His | p.Arg2336His | 9 | 13903 | 0.00065 | 9 | 13903 | 0.00065 | 1.00 | 0.00 | 0.999972397 | 0.999986988 |
| 97 | p.Arg2163His | p.Arg177Cys | 9 | 13903 | 0.00065 | 11 | 13904 | 0.00079 | 0.82 | 0.20 | 0.654615239 | 0.695261283 |
| 98 | p.Arg2163His | p.Arg2458His | 9 | 13903 | 0.00065 | 17 | 34758 | 0.00049 | 1.32 | 0.45 | 0.503233645 | 0.552563749 |
| 99 | p.Arg2163His | p.Arg2454His | 9 | 13903 | 0.00065 | 16 | 34757 | 0.00046 | 1.41 | 0.65 | 0.421553361 | 0.482436329 |

| | A | B | C | D | E | F | G | H | I | J | K | L |
|---|---|---|---|---|---|---|---|---|---|---|---|---|
| 100 | p.Arg2163His | p.Gly3990Val | 9 | 13903 | 0.00065 | 11 | 27806 | 0.00040 | 1.64 | 1.17 | 0.279454859 | 0.340179449 |
| 101 | p.Arg2163His | p.Val2168Met | 9 | 13903 | 0.00065 | 8 | 34757 | 0.00023 | 2.81 | 4.43 | 0.035411752 | 0.055335244 |
| 102 | p.Arg2163His | p.Gly248Arg | 9 | 13903 | 0.00065 | 5 | 27807 | 0.00018 | 3.60 | 5.58 | 0.018156343 | 0.031389828 |
| 103 | p.Arg2163His | p.Thr2206Met | 9 | 13903 | 0.00065 | 28 | 139031 | 0.00020 | 3.21 | 7.45 | 0.006360287 | 0.011949737 |
| 104 | p.Arg2163His | p.Gly2434Arg | 9 | 13903 | 0.00065 | 127 | 980149 | 0.00013 | 5.00 | 14.16 | 0.000167589 | 0.000453943 |
| 105 | p.Arg2163His | p.Glu3104Lys | 9 | 13903 | 0.00065 | 6 | 62563 | 0.00010 | 6.75 | 12.90 | 0.000328009 | 0.000813852 |
| 106 | p.Arg2163His | p.Arg4737Gln | 9 | 13903 | 0.00065 | 9 | 125128 | 0.00007 | 9.00 | 18.39 | 1.80E-05 | 5.63E-05 |
| 107 | p.Arg2163His | p.Arg2355Trp | 9 | 13903 | 0.00065 | 9 | 139029 | 0.00006 | 10.00 | 19.92 | 8.06E-06 | 2.82E-05 |
| 108 | p.Arg2163His | p.Val4849Ile | 9 | 13903 | 0.00065 | 8 | 145983 | 0.00005 | 11.81 | 21.91 | 2.86E-06 | 1.08E-05 |
| 109 | p.Arg2163His | p.Ser1728Phe | 9 | 13903 | 0.00065 | 9 | 430988 | 0.00002 | 31.00 | 38.00 | 7.07E-10 | 5.36E-09 |
| 110 | p.Arg2163His | p.Arg614Cys | 9 | 13903 | 0.00065 | 16 | 424097 | 0.00004 | 17.16 | 30.46 | 3.40E-08 | 1.94E-07 |
| 111 | p.Arg2163His | p.Asp3986Glu | 9 | 13903 | 0.00065 | 8 | 6951 | 0.00115 | 0.56 | 1.37 | 0.242160222 | 0.298113909 |
| 112 | p.Arg2163His | p.His2204Gln | 9 | 13903 | 0.00065 | 5 | 6952 | 0.00072 | 0.90 | 0.04 | 0.850990575 | 0.872466678 |
| 113 | p.Arg2163His | p.Arg552Trp | 9 | 13903 | 0.00065 | 5 | 20856 | 0.00024 | 2.70 | 3.35 | 0.067085523 | 0.097776968 |
| 114 | p.Arg2163His | p.Arg3772Gln | 9 | 13903 | 0.00065 | 7 | 20855 | 0.00034 | 1.93 | 1.71 | 0.190354073 | 0.244540064 |
| 115 | p.Arg2163His | p.Phe2364Val | 9 | 13903 | 0.00065 | 8 | 76478 | 0.00010 | 6.19 | 12.86 | 0.000335826 | 0.000820719 |
| 116 | p.Arg2163His | p.Pro4973Leu | 9 | 13903 | 0.00065 | 6 | 243302 | 0.00002 | 26.25 | 33.00 | 9.23E-09 | 5.73E-08 |
| 117 | p.Arg2336His | p.Arg177Cys | 9 | 13903 | 0.00065 | 11 | 13904 | 0.00079 | 0.82 | 0.20 | 0.654588935 | 0.695261283 |
| 118 | p.Arg2336His | p.Arg2458His | 9 | 13903 | 0.00065 | 17 | 34758 | 0.00049 | 1.32 | 0.45 | 0.503258061 | 0.552563749 |
| 119 | p.Arg2336His | p.Arg2454His | 9 | 13903 | 0.00065 | 16 | 34757 | 0.00046 | 1.41 | 0.65 | 0.42157513 | 0.482436329 |
| 120 | p.Arg2336His | p.Gly3990Val | 9 | 13903 | 0.00065 | 11 | 27806 | 0.00040 | 1.64 | 1.17 | 0.279470501 | 0.340179449 |
| 121 | p.Arg2336His | p.Val2168Met | 9 | 13903 | 0.00065 | 8 | 34757 | 0.00023 | 2.81 | 4.43 | 0.035414556 | 0.055335244 |
| 122 | p.Arg2336His | p.Gly248Arg | 9 | 13903 | 0.00065 | 5 | 27807 | 0.00018 | 3.60 | 5.58 | 0.018157808 | 0.031389828 |
| 123 | p.Arg2336His | p.Thr2206Met | 9 | 13903 | 0.00065 | 28 | 139031 | 0.00020 | 3.21 | 7.45 | 0.006360937 | 0.011949737 |
| 124 | p.Arg2336His | p.Gly2434Arg | 9 | 13903 | 0.00065 | 127 | 980149 | 0.00013 | 5.00 | 14.16 | 0.00016761 | 0.000453943 |
| 125 | p.Arg2336His | p.Glu3104Lys | 9 | 13903 | 0.00065 | 6 | 62563 | 0.00010 | 6.75 | 12.90 | 0.000328045 | 0.000813852 |
| 126 | p.Arg2336His | p.Arg4737Gln | 9 | 13903 | 0.00065 | 9 | 125128 | 0.00007 | 9.00 | 18.39 | 1.80E-05 | 5.63E-05 |
| 127 | p.Arg2336His | p.Arg2355Trp | 9 | 13903 | 0.00065 | 9 | 139029 | 0.00006 | 10.00 | 19.92 | 8.06E-06 | 2.82E-05 |
| 128 | p.Arg2336His | p.Val4849Ile | 9 | 13903 | 0.00065 | 8 | 145983 | 0.00005 | 11.81 | 21.91 | 2.86E-06 | 1.08E-05 |
| 129 | p.Arg2336His | p.Ser1728Phe | 9 | 13903 | 0.00065 | 9 | 430988 | 0.00002 | 31.00 | 38.00 | 7.07E-10 | 5.36E-09 |
| 130 | p.Arg2336His | p.Arg614Cys | 9 | 13903 | 0.00065 | 16 | 424097 | 0.00004 | 17.16 | 30.46 | 3.40E-08 | 1.94E-07 |
| 131 | p.Arg2336His | p.Asp3986Glu | 9 | 13903 | 0.00065 | 8 | 6951 | 0.00115 | 0.56 | 1.37 | 0.242147123 | 0.298113909 |
| 132 | p.Arg2336His | p.His2204Gln | 9 | 13903 | 0.00065 | 5 | 6952 | 0.00072 | 0.90 | 0.04 | 0.850967897 | 0.872466678 |

| | A | B | C | D | E | F | G | H | I | J | K | L |
|---|---|---|---|---|---|---|---|---|---|---|---|---|
| 133 | p.Arg2336His | p.Arg552Trp | 9 | 13903 | 0.00065 | 5 | 20856 | 0.00024 | 2.70 | 3.35 | 0.067090043 | 0.097776968 |
| 134 | p.Arg2336His | p.Arg3772Gln | 9 | 13903 | 0.00065 | 7 | 20855 | 0.00034 | 1.93 | 1.71 | 0.190365034 | 0.244540064 |
| 135 | p.Arg2336His | p.Phe2364Val | 9 | 13903 | 0.00065 | 8 | 76478 | 0.00010 | 6.19 | 12.86 | 0.000335864 | 0.000820719 |
| 136 | p.Arg2336His | p.Pro4973Leu | 9 | 13903 | 0.00065 | 6 | 243302 | 0.00002 | 26.25 | 33.00 | 9.23E-09 | 5.73E-08 |
| 137 | p.Arg177Cys | p.Arg2458His | 11 | 13904 | 0.00079 | 17 | 34758 | 0.00049 | 1.62 | 1.48 | 0.223803852 | 0.28083495 |
| 138 | p.Arg177Cys | p.Arg2454His | 11 | 13904 | 0.00079 | 16 | 34757 | 0.00046 | 1.72 | 1.83 | 0.17628687 | 0.229200591 |
| 139 | p.Arg177Cys | p.Gly3990Val | 11 | 13904 | 0.00079 | 11 | 27806 | 0.00040 | 2.00 | 2.59 | 0.107503738 | 0.148045402 |
| 140 | p.Arg177Cys | p.Val2168Met | 11 | 13904 | 0.00079 | 8 | 34757 | 0.00023 | 3.44 | 7.08 | 0.007797618 | 0.01448129 |
| 141 | p.Arg177Cys | p.Gly248Arg | 11 | 13904 | 0.00079 | 5 | 27807 | 0.00018 | 4.40 | 8.35 | 0.003859268 | 0.007735588 |
| 142 | p.Arg177Cys | p.Thr2206Met | 11 | 13904 | 0.00079 | 28 | 139031 | 0.00020 | 3.93 | 11.69 | 0.000628642 | 0.001480498 |
| 143 | p.Arg177Cys | p.Gly2434Arg | 11 | 13904 | 0.00079 | 127 | 980149 | 0.00013 | 6.11 | 20.76 | 5.19E-06 | 1.92E-05 |
| 144 | p.Arg177Cys | p.Glu3104Lys | 11 | 13904 | 0.00079 | 6 | 62563 | 0.00010 | 8.25 | 17.84 | 2.41E-05 | 7.24E-05 |
| 145 | p.Arg177Cys | p.Arg4737Gln | 11 | 13904 | 0.00079 | 9 | 125128 | 0.00007 | 11.00 | 25.03 | 5.66E-07 | 2.42E-06 |
| 146 | p.Arg177Cys | p.Arg2355Trp | 11 | 13904 | 0.00079 | 9 | 139029 | 0.00006 | 12.22 | 26.94 | 2.10E-07 | 1.02E-06 |
| 147 | p.Arg177Cys | p.Val4849Ile | 11 | 13904 | 0.00079 | 8 | 145983 | 0.00005 | 14.44 | 29.32 | 6.13E-08 | 3.18E-07 |
| 148 | p.Arg177Cys | p.Ser1728Phe | 11 | 13904 | 0.00079 | 9 | 430988 | 0.00002 | 37.89 | 49.29 | 2.21E-12 | 2.99E-11 |
| 149 | p.Arg177Cys | p.Arg614Cys | 11 | 13904 | 0.00079 | 16 | 424097 | 0.00004 | 20.97 | 40.43 | 2.03E-10 | 1.74E-09 |
| 150 | p.Arg177Cys | p.Asp3986Glu | 11 | 13904 | 0.00079 | 8 | 6951 | 0.00115 | 0.69 | 0.63 | 0.425740731 | 0.483796286 |
| 151 | p.Arg177Cys | p.His2204Gln | 11 | 13904 | 0.00079 | 5 | 6952 | 0.00072 | 1.10 | 0.03 | 0.859029769 | 0.87793923 |
| 152 | p.Arg177Cys | p.Arg552Trp | 11 | 13904 | 0.00079 | 5 | 20856 | 0.00024 | 3.30 | 5.39 | 0.020231654 | 0.034246289 |
| 153 | p.Arg177Cys | p.Arg3772Gln | 11 | 13904 | 0.00079 | 7 | 20855 | 0.00034 | 2.36 | 3.25 | 0.071317376 | 0.103013988 |
| 154 | p.Arg177Cys | p.Phe2364Val | 11 | 13904 | 0.00079 | 8 | 76478 | 0.00010 | 7.56 | 17.99 | 2.22E-05 | 6.75E-05 |
| 155 | p.Arg177Cys | p.Pro4973Leu | 11 | 13904 | 0.00079 | 6 | 243302 | 0.00002 | 32.08 | 42.78 | 6.12E-11 | 5.68E-10 |
| 156 | p.Arg2458His | p.Arg2454His | 17 | 34758 | 0.00049 | 16 | 34757 | 0.00046 | 1.06 | 0.03 | 0.861798562 | 0.878007939 |
| 157 | p.Arg2458His | p.Gly3990Val | 17 | 34758 | 0.00049 | 11 | 27806 | 0.00040 | 1.24 | 0.30 | 0.580956529 | 0.629369573 |
| 158 | p.Arg2458His | p.Val2168Met | 17 | 34758 | 0.00049 | 8 | 34757 | 0.00023 | 2.12 | 3.31 | 0.06869916 | 0.09967512 |
| 159 | p.Arg2458His | p.Gly248Arg | 17 | 34758 | 0.00049 | 5 | 27807 | 0.00018 | 2.72 | 4.51 | 0.033653097 | 0.053352471 |
| 160 | p.Arg2458His | p.Thr2206Met | 17 | 34758 | 0.00049 | 28 | 139031 | 0.00020 | 2.43 | 7.55 | 0.006000648 | 0.01140474 |
| 161 | p.Arg2458His | p.Gly2434Arg | 17 | 34758 | 0.00049 | 127 | 980149 | 0.00013 | 3.77 | 19.02 | 1.29E-05 | 4.31E-05 |
| 162 | p.Arg2458His | p.Glu3104Lys | 17 | 34758 | 0.00049 | 6 | 62563 | 0.00010 | 5.10 | 13.91 | 0.000192125 | 0.000511808 |
| 163 | p.Arg2458His | p.Arg4737Gln | 17 | 34758 | 0.00049 | 9 | 125128 | 0.00007 | 6.80 | 22.76 | 1.84E-06 | 7.29E-06 |
| 164 | p.Arg2458His | p.Arg2355Trp | 17 | 34758 | 0.00049 | 9 | 139029 | 0.00006 | 7.56 | 25.20 | 5.18E-07 | 2.24E-06 |
| 165 | p.Arg2458His | p.Val4849Ile | 17 | 34758 | 0.00049 | 8 | 145983 | 0.00005 | 8.93 | 28.13 | 1.14E-07 | 5.77E-07 |

| | A | B | C | D | E | F | G | H | I | J | K | L |
|---|---|---|---|---|---|---|---|---|---|---|---|---|
| 166 | p.Arg2458His | p.Ser1728Phe | 17 | 34758 | 0.00049 | 9 | 430988 | 0.00002 | 23.42 | 56.09 | 6.91E-14 | 1.18E-12 |
| 167 | p.Arg2458His | p.Arg614Cys | 17 | 34758 | 0.00049 | 16 | 424097 | 0.00004 | 12.96 | 44.53 | 2.50E-11 | 2.62E-10 |
| 168 | p.Arg2458His | p.Asp3986Glu | 17 | 34758 | 0.00049 | 8 | 6951 | 0.00115 | 0.42 | 3.52 | 0.060499338 | 0.090193967 |
| 169 | p.Arg2458His | p.His2204Gln | 17 | 34758 | 0.00049 | 5 | 6952 | 0.00072 | 0.68 | 0.53 | 0.464828217 | 0.520928174 |
| 170 | p.Arg2458His | p.Arg552Trp | 17 | 34758 | 0.00049 | 5 | 20856 | 0.00024 | 2.04 | 2.21 | 0.137413729 | 0.183030581 |
| 171 | p.Arg2458His | p.Arg3772Gln | 17 | 34758 | 0.00049 | 7 | 20855 | 0.00034 | 1.46 | 0.74 | 0.390561292 | 0.456591439 |
| 172 | p.Arg2458His | p.Phe2364Val | 17 | 34758 | 0.00049 | 8 | 76478 | 0.00010 | 4.68 | 14.20 | 0.000164247 | 0.000452376 |
| 173 | p.Arg2458His | p.Pro4973Leu | 17 | 34758 | 0.00049 | 6 | 243302 | 0.00002 | 19.83 | 45.90 | 1.24E-11 | 1.35E-10 |
| 174 | p.Arg2454His | p.Gly3990Val | 16 | 34757 | 0.00046 | 11 | 27806 | 0.00040 | 1.16 | 0.15 | 0.697606084 | 0.736110316 |
| 175 | p.Arg2454His | p.Val2168Met | 16 | 34757 | 0.00046 | 8 | 34757 | 0.00023 | 2.00 | 2.72 | 0.099200391 | 0.13777832 |
| 176 | p.Arg2454His | p.Gly248Arg | 16 | 34757 | 0.00046 | 5 | 27807 | 0.00018 | 2.56 | 3.87 | 0.049269193 | 0.074824709 |
| 177 | p.Arg2454His | p.Thr2206Met | 16 | 34757 | 0.00046 | 28 | 139031 | 0.00020 | 2.29 | 6.32 | 0.011966854 | 0.021137106 |
| 178 | p.Arg2454His | p.Gly2434Arg | 16 | 34757 | 0.00046 | 127 | 980149 | 0.00013 | 3.55 | 16.60 | 4.62E-05 | 0.000136572 |
| 179 | p.Arg2454His | p.Glu3104Lys | 16 | 34757 | 0.00046 | 6 | 62563 | 0.00010 | 4.80 | 12.47 | 0.00041402 | 0.000989386 |
| 180 | p.Arg2454His | p.Arg4737Gln | 16 | 34757 | 0.00046 | 9 | 125128 | 0.00007 | 6.40 | 20.58 | 5.73E-06 | 2.09E-05 |
| 181 | p.Arg2454His | p.Arg2355Trp | 16 | 34757 | 0.00046 | 9 | 139029 | 0.00006 | 7.11 | 22.85 | 1.75E-06 | 7.12E-06 |
| 182 | p.Arg2454His | p.Val4849Ile | 16 | 34757 | 0.00046 | 8 | 145983 | 0.00005 | 8.40 | 25.62 | 4.15E-07 | 1.82E-06 |
| 183 | p.Arg2454His | p.Ser1728Phe | 16 | 34757 | 0.00046 | 9 | 430988 | 0.00002 | 22.04 | 51.77 | 6.23E-13 | 9.64E-12 |
| 184 | p.Arg2454His | p.Arg614Cys | 16 | 34757 | 0.00046 | 16 | 424097 | 0.00004 | 12.20 | 40.73 | 1.75E-10 | 1.54E-09 |
| 185 | p.Arg2454His | p.Asp3986Glu | 16 | 34757 | 0.00046 | 8 | 6951 | 0.00115 | 0.40 | 3.95 | 0.046877564 | 0.0715268 |
| 186 | p.Arg2454His | p.His2204Gln | 16 | 34757 | 0.00046 | 5 | 6952 | 0.00072 | 0.64 | 0.70 | 0.40308654 | 0.464722551 |
| 187 | p.Arg2454His | p.Arg552Trp | 16 | 34757 | 0.00046 | 5 | 20856 | 0.00024 | 1.92 | 1.80 | 0.180179644 | 0.233300335 |
| 188 | p.Arg2454His | p.Arg3772Gln | 16 | 34757 | 0.00046 | 7 | 20855 | 0.00034 | 1.37 | 0.50 | 0.477477433 | 0.531438924 |
| 189 | p.Arg2454His | p.Phe2364Val | 16 | 34757 | 0.00046 | 8 | 76478 | 0.00010 | 4.40 | 12.67 | 0.000372418 | 0.000896561 |
| 190 | p.Arg2454His | p.Pro4973Leu | 16 | 34757 | 0.00046 | 6 | 243302 | 0.00002 | 18.67 | 42.36 | 7.58E-11 | 6.85E-10 |
| 191 | p.Gly3990Val | p.Val2168Met | 11 | 27806 | 0.00040 | 8 | 34757 | 0.00023 | 1.72 | 1.38 | 0.239908615 | 0.298113909 |
| 192 | p.Gly3990Val | p.Gly248Arg | 11 | 27806 | 0.00040 | 5 | 27807 | 0.00018 | 2.20 | 2.31 | 0.128854452 | 0.173766377 |
| 193 | p.Gly3990Val | p.Thr2206Met | 11 | 27806 | 0.00040 | 28 | 139031 | 0.00020 | 1.96 | 3.23 | 0.072381718 | 0.103630212 |
| 194 | p.Gly3990Val | p.Gly2434Arg | 11 | 27806 | 0.00040 | 127 | 980149 | 0.00013 | 3.05 | 9.35 | 0.002229725 | 0.004736345 |
| 195 | p.Gly3990Val | p.Glu3104Lys | 11 | 27806 | 0.00040 | 6 | 62563 | 0.00010 | 4.13 | 8.27 | 0.004033519 | 0.007993255 |
| 196 | p.Gly3990Val | p.Arg4737Gln | 11 | 27806 | 0.00040 | 9 | 125128 | 0.00007 | 5.50 | 13.59 | 0.00022725 | 0.000600457 |
| 197 | p.Gly3990Val | p.Arg2355Trp | 11 | 27806 | 0.00040 | 9 | 139029 | 0.00006 | 6.11 | 15.17 | 9.80E-05 | 0.000274564 |
| 198 | p.Gly3990Val | p.Val4849Ile | 11 | 27806 | 0.00040 | 8 | 145983 | 0.00005 | 7.22 | 17.24 | 3.29E-05 | 9.81E-05 |

| | A | B | C | D | E | F | G | H | I | J | K | L |
|---|---|---|---|---|---|---|---|---|---|---|---|---|
| 199 | p.Gly3990Val | p.Ser1728Phe | 11 | 27806 | 0.00040 | 9 | 430988 | 0.00002 | 18.94 | 35.27 | 2.86E-09 | 1.94E-08 |
| 200 | p.Gly3990Val | p.Arg614Cys | 11 | 27806 | 0.00040 | 16 | 424097 | 0.00004 | 10.49 | 26.87 | 2.17E-07 | 1.04E-06 |
| 201 | p.Gly3990Val | p.Asp3986Glu | 11 | 27806 | 0.00040 | 8 | 6951 | 0.00115 | 0.34 | 4.80 | 0.028522133 | 0.046348466 |
| 202 | p.Gly3990Val | p.His2204Gln | 11 | 27806 | 0.00040 | 5 | 6952 | 0.00072 | 0.55 | 1.13 | 0.288055961 | 0.348023002 |
| 203 | p.Gly3990Val | p.Arg552Trp | 11 | 27806 | 0.00040 | 5 | 20856 | 0.00024 | 1.65 | 0.91 | 0.3400952 | 0.401930691 |
| 204 | p.Gly3990Val | p.Arg3772Gln | 11 | 27806 | 0.00040 | 7 | 20855 | 0.00034 | 1.18 | 0.12 | 0.732533168 | 0.765508938 |
| 205 | p.Gly3990Val | p.Phe2364Val | 11 | 27806 | 0.00040 | 8 | 76478 | 0.00010 | 3.78 | 8.18 | 0.004237865 | 0.00834731 |
| 206 | p.Gly3990Val | p.Pro4973Leu | 11 | 27806 | 0.00040 | 6 | 243302 | 0.00002 | 16.04 | 29.32 | 6.12E-08 | 3.18E-07 |
| 207 | p.Val2168Met | p.Gly248Arg | 8 | 34757 | 0.00023 | 5 | 27807 | 0.00018 | 1.28 | 0.19 | 0.662310913 | 0.701143475 |
| 208 | p.Val2168Met | p.Thr2206Met | 8 | 34757 | 0.00023 | 28 | 139031 | 0.00020 | 1.14 | 0.11 | 0.742181293 | 0.773105513 |
| 209 | p.Val2168Met | p.Gly2434Arg | 8 | 34757 | 0.00023 | 127 | 980149 | 0.00013 | 1.78 | 2.11 | 0.146520327 | 0.193573602 |
| 210 | p.Val2168Met | p.Glu3104Lys | 8 | 34757 | 0.00023 | 6 | 62563 | 0.00010 | 2.40 | 2.65 | 0.103258729 | 0.142804625 |
| 211 | p.Val2168Met | p.Arg4737Gln | 8 | 34757 | 0.00023 | 9 | 125128 | 0.00007 | 3.20 | 5.32 | 0.021068933 | 0.035478774 |
| 212 | p.Val2168Met | p.Arg2355Trp | 8 | 34757 | 0.00023 | 9 | 139029 | 0.00006 | 3.56 | 6.26 | 0.012353559 | 0.021702198 |
| 213 | p.Val2168Met | p.Val4849Ile | 8 | 34757 | 0.00023 | 8 | 145983 | 0.00005 | 4.20 | 7.62 | 0.00578798 | 0.011065257 |
| 214 | p.Val2168Met | p.Ser1728Phe | 8 | 34757 | 0.00023 | 9 | 430988 | 0.00002 | 11.02 | 19.41 | 1.05E-05 | 3.60E-05 |
| 215 | p.Val2168Met | p.Arg614Cys | 8 | 34757 | 0.00023 | 16 | 424097 | 0.00004 | 6.10 | 13.25 | 0.000272076 | 0.000690819 |
| 216 | p.Val2168Met | p.Asp3986Glu | 8 | 34757 | 0.00023 | 8 | 6951 | 0.00115 | 0.20 | 9.40 | 0.002164459 | 0.004627955 |
| 217 | p.Val2168Met | p.His2204Gln | 8 | 34757 | 0.00023 | 5 | 6952 | 0.00072 | 0.32 | 3.51 | 0.06094935 | 0.090449948 |
| 218 | p.Val2168Met | p.Arg552Trp | 8 | 34757 | 0.00023 | 5 | 20856 | 0.00024 | 0.96 | 0.01 | 0.94312256 | 0.95403047 |
| 219 | p.Val2168Met | p.Arg3772Gln | 8 | 34757 | 0.00023 | 7 | 20855 | 0.00034 | 0.69 | 0.52 | 0.469175195 | 0.523992916 |
| 220 | p.Val2168Met | p.Phe2364Val | 8 | 34757 | 0.00023 | 8 | 76478 | 0.00010 | 2.20 | 2.43 | 0.119359636 | 0.162309129 |
| 221 | p.Val2168Met | p.Pro4973Leu | 8 | 34757 | 0.00023 | 6 | 243302 | 0.00002 | 9.33 | 15.75 | 7.22E-05 | 0.000209538 |
| 222 | p.Gly248Arg | p.Thr2206Met | 5 | 27807 | 0.00018 | 28 | 139031 | 0.00020 | 0.89 | 0.06 | 0.812977465 | 0.841457568 |
| 223 | p.Gly248Arg | p.Gly2434Arg | 5 | 27807 | 0.00018 | 127 | 980149 | 0.00013 | 1.39 | 0.47 | 0.494008807 | 0.546324623 |
| 224 | p.Gly248Arg | p.Glu3104Lys | 5 | 27807 | 0.00018 | 6 | 62563 | 0.00010 | 1.87 | 1.04 | 0.307617028 | 0.36891341 |
| 225 | p.Gly248Arg | p.Arg4737Gln | 5 | 27807 | 0.00018 | 9 | 125128 | 0.00007 | 2.50 | 2.41 | 0.120547766 | 0.163241766 |
| 226 | p.Gly248Arg | p.Arg2355Trp | 5 | 27807 | 0.00018 | 9 | 139029 | 0.00006 | 2.78 | 2.95 | 0.085882583 | 0.120309652 |
| 227 | p.Gly248Arg | p.Val4849Ile | 5 | 27807 | 0.00018 | 8 | 145983 | 0.00005 | 3.28 | 3.79 | 0.051497618 | 0.077845236 |
| 228 | p.Gly248Arg | p.Ser1728Phe | 5 | 27807 | 0.00018 | 9 | 430988 | 0.00002 | 8.61 | 10.91 | 0.00095682 | 0.00215949 |
| 229 | p.Gly248Arg | p.Arg614Cys | 5 | 27807 | 0.00018 | 16 | 424097 | 0.00004 | 4.77 | 6.86 | 0.008808398 | 0.015992901 |
| 230 | p.Gly248Arg | p.Asp3986Glu | 5 | 27807 | 0.00018 | 8 | 6951 | 0.00115 | 0.16 | 10.66 | 0.001094933 | 0.002437351 |
| 231 | p.Gly248Arg | p.His2204Gln | 5 | 27807 | 0.00018 | 5 | 6952 | 0.00072 | 0.25 | 4.46 | 0.034636006 | 0.054644185 |

| | A | B | C | D | E | F | G | H | I | J | K | L |
|---|---|---|---|---|---|---|---|---|---|---|---|---|
| 232 | p.Gly248Arg | p.Arg552Trp | 5 | 27807 | 0.00018 | 5 | 20856 | 0.00024 | 0.75 | 0.21 | 0.649804829 | 0.694692662 |
| 233 | p.Gly248Arg | p.Arg3772Gln | 5 | 27807 | 0.00018 | 7 | 20855 | 0.00034 | 0.54 | 1.16 | 0.281920046 | 0.341880653 |
| 234 | p.Gly248Arg | p.Phe2364Val | 5 | 27807 | 0.00018 | 8 | 76478 | 0.00010 | 1.72 | 0.86 | 0.35455035 | 0.417495884 |
| 235 | p.Gly248Arg | p.Pro4973Leu | 5 | 27807 | 0.00018 | 6 | 243302 | 0.00002 | 7.29 | 8.91 | 0.002832014 | 0.005825346 |
| 236 | p.Thr2206Met | p.Gly2434Arg | 28 | 139031 | 0.00020 | 127 | 980149 | 0.00013 | 1.55 | 4.05 | 0.044062547 | 0.068192037 |
| 237 | p.Thr2206Met | p.Glu3104Lys | 28 | 139031 | 0.00020 | 6 | 62563 | 0.00010 | 2.10 | 3.16 | 0.07544685 | 0.107544853 |
| 238 | p.Thr2206Met | p.Arg4737Gln | 28 | 139031 | 0.00020 | 9 | 125128 | 0.00007 | 2.80 | 8.34 | 0.003879695 | 0.007735588 |
| 239 | p.Thr2206Met | p.Arg2355Trp | 28 | 139031 | 0.00020 | 9 | 139029 | 0.00006 | 3.11 | 10.24 | 0.001375654 | 0.003020862 |
| 240 | p.Thr2206Met | p.Val4849Ile | 28 | 139031 | 0.00020 | 8 | 145983 | 0.00005 | 3.68 | 12.77 | 0.000353162 | 0.00085655 |
| 241 | p.Thr2206Met | p.Ser1728Phe | 28 | 139031 | 0.00020 | 9 | 430988 | 0.00002 | 9.64 | 42.99 | 5.49E-11 | 5.25E-10 |
| 242 | p.Thr2206Met | p.Arg614Cys | 28 | 139031 | 0.00020 | 16 | 424097 | 0.00004 | 5.34 | 29.72 | 4.98E-08 | 2.74E-07 |
| 243 | p.Thr2206Met | p.Asp3986Glu | 28 | 139031 | 0.00020 | 8 | 6951 | 0.00115 | 0.17 | 13.31 | 0.000264574 | 0.000682432 |
| 244 | p.Thr2206Met | p.His2204Gln | 28 | 139031 | 0.00020 | 5 | 6952 | 0.00072 | 0.28 | 5.11 | 0.023847047 | 0.039745079 |
| 245 | p.Thr2206Met | p.Arg552Trp | 28 | 139031 | 0.00020 | 5 | 20856 | 0.00024 | 0.84 | 0.12 | 0.725032832 | 0.762574985 |
| 246 | p.Thr2206Met | p.Arg3772Gln | 28 | 139031 | 0.00020 | 7 | 20855 | 0.00034 | 0.60 | 1.31 | 0.251524481 | 0.30847342 |
| 247 | p.Thr2206Met | p.Phe2364Val | 28 | 139031 | 0.00020 | 8 | 76478 | 0.00010 | 1.93 | 2.98 | 0.084183493 | 0.11843998 |
| 248 | p.Thr2206Met | p.Pro4973Leu | 28 | 139031 | 0.00020 | 6 | 243302 | 0.00002 | 8.17 | 30.39 | 3.54E-08 | 1.98E-07 |
| 249 | p.Gly2434Arg | p.Glu3104Lys | 127 | 980149 | 0.00013 | 6 | 62563 | 0.00010 | 1.35 | 0.57 | 0.450625161 | 0.508517977 |
| 250 | p.Gly2434Arg | p.Arg4737Gln | 127 | 980149 | 0.00013 | 9 | 125128 | 0.00007 | 1.80 | 3.46 | 0.062795856 | 0.092766606 |
| 251 | p.Gly2434Arg | p.Arg2355Trp | 127 | 980149 | 0.00013 | 9 | 139029 | 0.00006 | 2.00 | 4.97 | 0.025860948 | 0.042881675 |
| 252 | p.Gly2434Arg | p.Val4849Ile | 127 | 980149 | 0.00013 | 8 | 145983 | 0.00005 | 2.36 | 7.22 | 0.007191681 | 0.013432737 |
| 253 | p.Gly2434Arg | p.Ser1728Phe | 127 | 980149 | 0.00013 | 9 | 430988 | 0.00002 | 6.20 | 47.65 | 5.09E-12 | 6.37E-11 |
| 254 | p.Gly2434Arg | p.Arg614Cys | 127 | 980149 | 0.00013 | 16 | 424097 | 0.00004 | 3.43 | 29.41 | 5.85E-08 | 3.17E-07 |
| 255 | p.Gly2434Arg | p.Asp3986Glu | 127 | 980149 | 0.00013 | 8 | 6951 | 0.00115 | 0.11 | 20.36 | 6.42E-06 | 2.29E-05 |
| 256 | p.Gly2434Arg | p.His2204Gln | 127 | 980149 | 0.00013 | 5 | 6952 | 0.00072 | 0.18 | 8.81 | 0.002993777 | 0.00608111 |
| 257 | p.Gly2434Arg | p.Arg552Trp | 127 | 980149 | 0.00013 | 5 | 20856 | 0.00024 | 0.54 | 1.52 | 0.218020235 | 0.274637893 |
| 258 | p.Gly2434Arg | p.Arg3772Gln | 127 | 980149 | 0.00013 | 7 | 20855 | 0.00034 | 0.39 | 4.59 | 0.032171162 | 0.051253076 |
| 259 | p.Gly2434Arg | p.Phe2364Val | 127 | 980149 | 0.00013 | 8 | 76478 | 0.00010 | 1.24 | 0.37 | 0.544445931 | 0.595774167 |
| 260 | p.Gly2434Arg | p.Pro4973Leu | 127 | 980149 | 0.00013 | 6 | 243302 | 0.00002 | 5.25 | 26.79 | 2.27E-07 | 1.07E-06 |
| 261 | p.Glu3104Lys | p.Arg4737Gln | 6 | 62563 | 0.00010 | 9 | 125128 | 0.00007 | 1.33 | 0.29 | 0.589326087 | 0.636315542 |
| 262 | p.Glu3104Lys | p.Arg2355Trp | 6 | 62563 | 0.00010 | 9 | 139029 | 0.00006 | 1.48 | 0.54 | 0.462997702 | 0.520672157 |
| 263 | p.Glu3104Lys | p.Val4849Ile | 6 | 62563 | 0.00010 | 8 | 145983 | 0.00005 | 1.75 | 1.03 | 0.309432334 | 0.369726134 |
| 264 | p.Glu3104Lys | p.Ser1728Phe | 6 | 62563 | 0.00010 | 9 | 430988 | 0.00002 | 4.59 | 7.03 | 0.007993713 | 0.014677721 |

| | A | B | C | D | E | F | G | H | I | J | K | L |
|---|---|---|---|---|---|---|---|---|---|---|---|---|
| 265 | p.Glu3104Lys | p.Arg614Cys | 6 | 62563 | 0.00010 | 16 | 424097 | 0.00004 | 2.54 | 3.24 | 0.07194476 | 0.103460385 |
| 266 | p.Glu3104Lys | p.Asp3986Glu | 6 | 62563 | 0.00010 | 8 | 6951 | 0.00115 | 0.08 | 18.98 | 1.32E-05 | 4.31E-05 |
| 267 | p.Glu3104Lys | p.His2204Gln | 6 | 62563 | 0.00010 | 5 | 6952 | 0.00072 | 0.13 | 9.13 | 0.002512068 | 0.005301442 |
| 268 | p.Glu3104Lys | p.Arg552Trp | 6 | 62563 | 0.00010 | 5 | 20856 | 0.00024 | 0.40 | 2.16 | 0.141963201 | 0.188318531 |
| 269 | p.Glu3104Lys | p.Arg3772Gln | 6 | 62563 | 0.00010 | 7 | 20855 | 0.00034 | 0.29 | 4.92 | 0.026619212 | 0.043914944 |
| 270 | p.Glu3104Lys | p.Phe2364Val | 6 | 62563 | 0.00010 | 8 | 76478 | 0.00010 | 0.92 | 0.03 | 0.871980947 | 0.88560565 |
| 271 | p.Glu3104Lys | p.Pro4973Leu | 6 | 62563 | 0.00010 | 6 | 243302 | 0.00002 | 3.89 | 5.15 | 0.023190728 | 0.038850446 |
| 272 | p.Arg4737Gln | p.Arg2355Trp | 9 | 125128 | 0.00007 | 9 | 139029 | 0.00006 | 1.11 | 0.05 | 0.8232142 | 0.849347984 |
| 273 | p.Arg4737Gln | p.Val4849Ile | 9 | 125128 | 0.00007 | 8 | 145983 | 0.00005 | 1.31 | 0.31 | 0.575297593 | 0.625323471 |
| 274 | p.Arg4737Gln | p.Ser1728Phe | 9 | 125128 | 0.00007 | 9 | 430988 | 0.00002 | 3.44 | 6.48 | 0.010882845 | 0.019433652 |
| 275 | p.Arg4737Gln | p.Arg614Cys | 9 | 125128 | 0.00007 | 16 | 424097 | 0.00004 | 1.91 | 2.23 | 0.135561127 | 0.182055232 |
| 276 | p.Arg4737Gln | p.Asp3986Glu | 9 | 125128 | 0.00007 | 8 | 6951 | 0.00115 | 0.06 | 24.58 | 7.14E-07 | 2.98E-06 |
| 277 | p.Arg4737Gln | p.His2204Gln | 9 | 125128 | 0.00007 | 5 | 6952 | 0.00072 | 0.10 | 12.17 | 0.000486078 | 0.001153105 |
| 278 | p.Arg4737Gln | p.Arg552Trp | 9 | 125128 | 0.00007 | 5 | 20856 | 0.00024 | 0.30 | 3.98 | 0.045928521 | 0.070742982 |
| 279 | p.Arg4737Gln | p.Arg3772Gln | 9 | 125128 | 0.00007 | 7 | 20855 | 0.00034 | 0.21 | 8.09 | 0.004457502 | 0.008727036 |
| 280 | p.Arg4737Gln | p.Phe2364Val | 9 | 125128 | 0.00007 | 8 | 76478 | 0.00010 | 0.69 | 0.59 | 0.443786729 | 0.502545948 |
| 281 | p.Arg4737Gln | p.Pro4973Leu | 9 | 125128 | 0.00007 | 6 | 243302 | 0.00002 | 2.92 | 4.23 | 0.039775309 | 0.061851558 |
| 282 | p.Arg2355Trp | p.Val4849Ile | 9 | 139029 | 0.00006 | 8 | 145983 | 0.00005 | 1.18 | 0.12 | 0.73144426 | 0.765508938 |
| 283 | p.Arg2355Trp | p.Ser1728Phe | 9 | 139029 | 0.00006 | 9 | 430988 | 0.00002 | 3.10 | 5.48 | 0.019268568 | 0.03278683 |
| 284 | p.Arg2355Trp | p.Arg614Cys | 9 | 139029 | 0.00006 | 16 | 424097 | 0.00004 | 1.72 | 1.58 | 0.208588086 | 0.263778708 |
| 285 | p.Arg2355Trp | p.Asp3986Glu | 9 | 139029 | 0.00006 | 8 | 6951 | 0.00115 | 0.06 | 26.08 | 3.27E-07 | 1.46E-06 |
| 286 | p.Arg2355Trp | p.His2204Gln | 9 | 139029 | 0.00006 | 5 | 6952 | 0.00072 | 0.09 | 13.07 | 0.00029942 | 0.000754354 |
| 287 | p.Arg2355Trp | p.Arg552Trp | 9 | 139029 | 0.00006 | 5 | 20856 | 0.00024 | 0.27 | 4.63 | 0.031329904 | 0.050407024 |
| 288 | p.Arg2355Trp | p.Arg3772Gln | 9 | 139029 | 0.00006 | 7 | 20855 | 0.00034 | 0.19 | 9.10 | 0.002553599 | 0.00535432 |
| 289 | p.Arg2355Trp | p.Phe2364Val | 9 | 139029 | 0.00006 | 8 | 76478 | 0.00010 | 0.62 | 0.96 | 0.327867234 | 0.388893616 |
| 290 | p.Arg2355Trp | p.Pro4973Leu | 9 | 139029 | 0.00006 | 6 | 243302 | 0.00002 | 2.63 | 3.44 | 0.063545969 | 0.093449955 |
| 291 | p.Val4849Ile | p.Ser1728Phe | 8 | 145983 | 0.00005 | 9 | 430988 | 0.00002 | 2.62 | 3.73 | 0.053395253 | 0.08034008 |
| 292 | p.Val4849Ile | p.Arg614Cys | 8 | 145983 | 0.00005 | 16 | 424097 | 0.00004 | 1.45 | 0.71 | 0.399452214 | 0.464722551 |
| 293 | p.Val4849Ile | p.Asp3986Glu | 8 | 145983 | 0.00005 | 8 | 6951 | 0.00115 | 0.05 | 28.02 | 1.20E-07 | 5.91E-07 |
| 294 | p.Val4849Ile | p.His2204Gln | 8 | 145983 | 0.00005 | 5 | 6952 | 0.00072 | 0.08 | 14.33 | 0.000153285 | 0.000425793 |
| 295 | p.Val4849Ile | p.Arg552Trp | 8 | 145983 | 0.00005 | 5 | 20856 | 0.00024 | 0.23 | 5.61 | 0.017886717 | 0.031253672 |
| 296 | p.Val4849Ile | p.Arg3772Gln | 8 | 145983 | 0.00005 | 7 | 20855 | 0.00034 | 0.16 | 10.52 | 0.001180293 | 0.00260949 |
| 297 | p.Val4849Ile | p.Phe2364Val | 8 | 145983 | 0.00005 | 8 | 76478 | 0.00010 | 0.52 | 1.64 | 0.199849646 | 0.254710333 |

| | A | B | C | D | E | F | G | H | I | J | K | L |
|---|---|---|---|---|---|---|---|---|---|---|---|---|
| 298 | p.Val4849Ile | p.Pro4973Leu | 8 | 145983 | 0.00005 | 6 | 243302 | 0.00002 | 2.22 | 2.21 | 0.136957242 | 0.183030581 |
| 299 | p.Ser1728Phe | p.Arg614Cys | 9 | 430988 | 0.00002 | 16 | 424097 | 0.00004 | 0.55 | 2.10 | 0.147212695 | 0.193700914 |
| 300 | p.Ser1728Phe | p.Asp3986Glu | 9 | 430988 | 0.00002 | 8 | 6951 | 0.00115 | 0.02 | 43.07 | 5.28E-11 | 5.20E-10 |
| 301 | p.Ser1728Phe | p.His2204Gln | 9 | 430988 | 0.00002 | 5 | 6952 | 0.00072 | 0.03 | 23.47 | 1.27E-06 | 5.22E-06 |
| 302 | p.Ser1728Phe | p.Arg552Trp | 9 | 430988 | 0.00002 | 5 | 20856 | 0.00024 | 0.09 | 13.36 | 0.000257276 | 0.000674313 |
| 303 | p.Ser1728Phe | p.Arg3772Gln | 9 | 430988 | 0.00002 | 7 | 20855 | 0.00034 | 0.06 | 21.98 | 2.75E-06 | 1.07E-05 |
| 304 | p.Ser1728Phe | p.Phe2364Val | 9 | 430988 | 0.00002 | 8 | 76478 | 0.00010 | 0.20 | 9.71 | 0.00183166 | 0.003968596 |
| 305 | p.Ser1728Phe | p.Pro4973Leu | 9 | 430988 | 0.00002 | 6 | 243302 | 0.00002 | 0.85 | 0.10 | 0.753789579 | 0.78268886 |
| 306 | p.Arg614Cys | p.Asp3986Glu | 16 | 424097 | 0.00004 | 8 | 6951 | 0.00115 | 0.03 | 36.00 | 1.97E-09 | 1.39E-08 |
| 307 | p.Arg614Cys | p.His2204Gln | 16 | 424097 | 0.00004 | 5 | 6952 | 0.00072 | 0.05 | 18.74 | 1.50E-05 | 4.82E-05 |
| 308 | p.Arg614Cys | p.Arg552Trp | 16 | 424097 | 0.00004 | 5 | 20856 | 0.00024 | 0.16 | 9.09 | 0.002574744 | 0.00536405 |
| 309 | p.Arg614Cys | p.Arg3772Gln | 16 | 424097 | 0.00004 | 7 | 20855 | 0.00034 | 0.11 | 16.11 | 5.96E-05 | 0.000174595 |
| 310 | p.Arg614Cys | p.Phe2364Val | 16 | 424097 | 0.00004 | 8 | 76478 | 0.00010 | 0.36 | 4.81 | 0.028247767 | 0.046348466 |
| 311 | p.Arg614Cys | p.Pro4973Leu | 16 | 424097 | 0.00004 | 6 | 243302 | 0.00002 | 1.53 | 0.84 | 0.360336225 | 0.42277716 |
| 312 | p.Asp3986Glu | p.His2204Gln | 8 | 6951 | 0.00115 | 5 | 6952 | 0.00072 | 1.60 | 0.70 | 0.403236183 | 0.464722551 |
| 313 | p.Asp3986Glu | p.Arg552Trp | 8 | 6951 | 0.00115 | 5 | 20856 | 0.00024 | 4.80 | 7.74 | 0.005415859 | 0.010415114 |
| 314 | p.Asp3986Glu | p.Arg3772Gln | 8 | 6951 | 0.00115 | 7 | 20855 | 0.00034 | 3.43 | 5.48 | 0.019227578 | 0.03278683 |
| 315 | p.Asp3986Glu | p.Phe2364Val | 8 | 6951 | 0.00115 | 8 | 76478 | 0.00010 | 11.00 | 18.97 | 1.33E-05 | 4.31E-05 |
| 316 | p.Asp3986Glu | p.Pro4973Leu | 8 | 6951 | 0.00115 | 6 | 243302 | 0.00002 | 46.67 | 38.55 | 5.33E-10 | 4.33E-09 |
| 317 | p.His2204Gln | p.Arg552Trp | 5 | 6952 | 0.00072 | 5 | 20856 | 0.00024 | 3.00 | 2.88 | 0.089846788 | 0.125322774 |
| 318 | p.His2204Gln | p.Arg3772Gln | 5 | 6952 | 0.00072 | 7 | 20855 | 0.00034 | 2.14 | 1.59 | 0.207350455 | 0.263237883 |
| 319 | p.His2204Gln | p.Phe2364Val | 5 | 6952 | 0.00072 | 8 | 76478 | 0.00010 | 6.88 | 8.92 | 0.002822191 | 0.005825346 |
| 320 | p.His2204Gln | p.Pro4973Leu | 5 | 6952 | 0.00072 | 6 | 243302 | 0.00002 | 29.17 | 21.01 | 4.56E-06 | 1.70E-05 |
| 321 | p.Arg552Trp | p.Arg3772Gln | 5 | 20856 | 0.00024 | 7 | 20855 | 0.00034 | 0.71 | 0.34 | 0.562715389 | 0.613699669 |
| 322 | p.Arg552Trp | p.Phe2364Val | 5 | 20856 | 0.00024 | 8 | 76478 | 0.00010 | 2.29 | 1.94 | 0.163657483 | 0.214470492 |
| 323 | p.Arg552Trp | p.Pro4973Leu | 5 | 20856 | 0.00024 | 6 | 243302 | 0.00002 | 9.72 | 11.22 | 0.0008102 | 0.001867483 |
| 324 | p.Arg3772Gln | p.Phe2364Val | 7 | 20855 | 0.00034 | 8 | 76478 | 0.00010 | 3.21 | 4.70 | 0.030195496 | 0.048823564 |
| 325 | p.Arg3772Gln | p.Pro4973Leu | 7 | 20855 | 0.00034 | 6 | 243302 | 0.00002 | 13.61 | 18.59 | 1.62E-05 | 5.17E-05 |
| 326 | p.Phe2364Val | p.Pro4973Leu | 8 | 76478 | 0.00010 | 6 | 243302 | 0.00002 | 4.24 | 7.05 | 0.007932517 | 0.014648114 |
