## Supplementary Tables 1 2 3 for "Epidemiology of malignant hyperthermia in the UK 1988-2025: implications for prevalence, mode of inheritance, relative risk associated with *RYR1* genotypes and *in vitro* contracture test phenotype"

Supplementary table S1. The number of MH reactions in the UK and number of referrals to the UK Malignant Hyperthermia Unit 1988-2025

| Year | No. of reactions | No. of referrals |
|---|---|---|
| 1988 | 24 | 20 |
| 1989 | 27 | 22 |
| 1990 | 19 | 16 |
| 1991 | 20 | 33 |
| 1992 | 34 | 29 |
| 1993 | 26 | 29 |
| 1994 | 16 | 18 |
| 1995 | 17 | 14 |
| 1996 | 14 | 25 |
| 1997 | 10 | 13 |
| 1998 | 12 | 11 |
| 1999 | 12 | 16 |
| 2000 | 16 | 18 |
| 2001 | 9 | 13 |
| 2002 | 6 | 9 |
| 2003 | 13 | 12 |
| 2004 | 8 | 15 |
| 2005 | 5 | 10 |
| 2006 | 9 | 10 |
| 2007 | 11 | 19 |
| 2008 | 6 | 13 |
| 2009 | 9 | 11 |
| 2010 | 11 | 18 |
| 2011 | 11 | 8 |
| 2012 | 8 | 19 |
| 2013 | 11 | 15 |
| 2014 | 8 | 12 |
| 2015 | 11 | 18 |
| 2016 | 9 | 14 |
| 2017 | 8 | 18 |
| 2018 | 9 | 18 |
| 2019 | 8 | 15 |
| 2020 | 7 | 14 |
| 2021 | 7 | 7 |
| 2022 | 3 | 2 |
| 2023 | 2 | 4 |
| 2024 | 0 | 0 |
| 2025 | 3 | 3 |

Supplementary table S2. Proportion of male and female MH index cases reacting with each general anaesthesia exposure

| GA exposure | Males | | | Females | | |
|---|---|---|---|---|---|---|
| | No. exposed | No. reacting | % reacting | No. exposed | No. reacting | % reacting |
| 1st | 247 | 101 | 41 | 128 | 60 | 47 |
| 2nd | 146 | 66 | 45 | 68 | 31 | 46 |
| 3rd | 80 | 40 | 50 | 37 | 20 | 54 |
| 4th | 40 | 26 | 65 | 17 | 6 | 35 |
| 5th | 14 | 9 | 64 | 11 | 7 | 64 |
| 6th | 5 | 3 | 60 | 4 | 2 | 50 |

Supplementary table S3. RYR1 variant allele frequencies in the UK Biobank whole genome sequence cohort and the non-UK Biobank subset of the non-Finnish European cohort of gnomAd (version 4.1)

| Variant ID | Transcript Consequence | Protein Consequence | UKB Allele count | UKB allele number | UKB AF | UKB 95% CI | Non-UKB NFE allele count in gnomAD | Non-UKB NFE allele number in gnomAD | Non-UKB AF | Non-UKB 95% CI |
|---|---|---|---|---|---|---|---|---|---|---|
| 19-38444211-C-T | c.487C>T | p.Arg163Cys | 1 | 980852 | 1.02E-06 | [1.801e-07, 5.780e-06] | 1 | 350094 | 2.86E-06 | [5.042e-07, 1.618e-05] |
| 19-38499997-G-A | c.7304G>A | p.Arg2435His | 0 | 980100 | 0 | [0.000e+00, 3.920e-06] | 1 | 350084 | 2.86E-06 | [5.042e-07, 1.618e-05] |
| 19-38448712-G-A | c.1021G>A | p.Gly341Arg | 5 | 981098 | 5.10E-06 | [2.177e-06, 1.193e-05] | 1 | 350106 | 2.86E-06 | [5.042e-07, 1.618e-05] |
| 19-38499655-G-A | c.7048G>A | p.Ala2350Thr | 1 | 981094 | 1.02E-06 | [1.799e-07, 5.774e-06] | 0 | 349992 | 0 | [0.000e+00, 1.098e-05] |
| 19-38580094-C-T | c.14477C>T | p.Thr4826Ile | 1 | 981084 | 1.02E-06 | [1.799e-07, 5.774e-06] | 1 | 350108 | 2.86E-06 | [5.042e-07, 1.618e-05] |
| 19-38494565-G-A | c.6488G>A | p.Arg2163His | 2 | 981096 | 2.04E-06 | [5.590e-07, 7.434e-06] | 0 | 350104 | 0 | [0.000e+00, 1.097e-05] |
| 19-38499223-G-A | c.7007G>A | p.Arg2336His | 2 | 981080 | 2.04E-06 | [5.590e-07, 7.434e-06] | 1 | 418138 | 2.39E-06 | [4.222e-07, 1.355e-05] |
| 19-38444253-C-T | c.529C>T | p.Arg177Cys | 2 | 980996 | 2.04E-06 | [5.591e-07, 7.434e-06] | 1 | 418034 | 2.39E-06 | [4.223e-07, 1.355e-05] |
| 19-38500655-G-A | c.7373G>A | p.Arg2458His | 5 | 981082 | 5.10E-06 | [2.177e-06, 1.193e-05] | 4 | 350104 | 1.14E-05 | [4.443e-06, 2.938e-05] |
| 19-38500643-G-A | c.7361G>A | p.Arg2454His | 5 | 981084 | 5.10E-06 | [2.177e-06, 1.193e-05] | 5 | 350104 | 1.43E-05 | [6.100e-06, 3.344e-05] |
| 19-38543832-G-T | c.11969G>T | p.Gly3990Val | 4 | 981088 | 4.08E-06 | [1.585e-06, 1.048e-05] | 0 | 350108 | 0 | [0.000e+00, 1.097e-05] |
| 19-38494579-G-A | c.6502G>A | p.Val2168Met | 5 | 981088 | 5.10E-06 | [2.177e-06, 1.193e-05] | 0 | 350104 | 0 | [0.000e+00, 1.097e-05] |
| 19-38446710-G-A | c.742G>A | p.Gly248Arg | 4 | 981038 | 4.08E-06 | [1.586e-06, 1.048e-05] | 1 | 350052 | 2.86E-06 | [5.043e-07, 1.618e-05] |
| 19-38496283-C-T | c.6617C>T | p.Thr2206Met | 20 | 981076 | 2.04E-05 | [1.320e-05, 3.149e-05] | 16 | 418144 | 3.83E-05 | [2.355e-05, 6.216e-05] |
| 19-38499993-G-A | c.7300G>A | p.Gly2434Arg | 141 | 981096 | 0.0001437 | [1.219e-04, 1.695e-04] | 32 | 418118 | 7.65E-05 | [5.422e-05, 1.080e-04] |
| 19-38512321-G-A | c.9310G>A | p.Glu3104Lys | 9 | 981088 | 9.17E-06 | [4.826e-06, 1.744e-05] | 2 | 350108 | 5.71E-06 | [1.567e-06, 2.083e-05] |
| 19-38577955-G-A | c.14210G>A | p.Arg4737Gln | 18 | 981074 | 1.84E-05 | [1.161e-05, 2.900e-05] | 2 | 350108 | 5.71E-06 | [1.567e-06, 2.083e-05] |
| 19-38499670-C-T | c.7063C>T | p.Arg2355Trp | 20 | 981092 | 2.04E-05 | [1.320e-05, 3.149e-05] | 7 | 417964 | 1.68E-05 | [8.113e-06, 3.457e-05] |
| 19-38580403-G-A | c.14545G>A | p.Val4849Ile | 21 | 981074 | 2.14E-05 | [1.400e-05, 3.272e-05] | 0 | 418130 | 0 | [0.000e+00, 9.187e-06] |
| 19-38485838-C-T | c.5183C>T | p.Ser1728Phe | 62 | 981094 | 6.32E-05 | [4.930e-05, 8.100e-05] | 6 | 418116 | 1.44E-05 | [6.577e-06, 3.131e-05] |
| 19-38457545-C-T | c.1840C>T | p.Arg614Cys | 61 | 980954 | 6.22E-05 | [4.842e-05, 7.987e-05] | 53 | 418128 | 0.0001268 | [9.692e-05, 1.658e-04] |
| 19-38543821-C-G | c.11958C>G | p.Asp3986Glu | 1 | 981086 | 1.02E-06 | [1.799e-07, 5.774e-06] | 1 | 350102 | 2.86E-06 | [5.042e-07, 1.618e-05] |
| 19-38496278-C-G | c.6612C>G | p.His2204Gln | 1 | 981064 | 1.02E-06 | [1.799e-07, 5.774e-06] | 2 | 350108 | 5.71E-06 | [1.567e-06, 2.083e-05] |
| 19-38455528-C-T | c.1654C>T | p.Arg552Trp | 3 | 981006 | 3.06E-06 | [1.040e-06, 8.992e-06] | 0 | 350104 | 0 | [0.000e+00, 1.097e-05] |

| | | | | | | | | | | |
|---|---|---|---|---|---|---|---|---|---|---|
| 19-38534775-G-A | c.11315G>A | p.Arg3772Gln | 3 | 981070 | 3.06E-06 | [1.040e-06, 8.992e-06] | 3 | 492462 | 6.09E-06 | [2.072e-06, 1.791e-05] |
| 19-38499697-T-G | c.7090T>G | p.Phe2364Val | 11 | 980940 | 1.12E-05 | [6.262e-06, 2.008e-05] | 0 | 349992 | 0 | [0.000e+00, 1.098e-05] |
| 19-38586140-C-T | c.14918C>T | p.Pro4973Leu | 35 | 981086 | 3.57E-05 | [2.565e-05, 4.961e-05] | 13 | 418130 | 3.11E-05 | [1.817e-05, 5.320e-05] |
| 19-38565034-G-C | c.12700G>C | p.Val4234Leu | 0 | 981090 | 0 | [0.000e+00, 3.916e-06] | 0 | 417964 | 0 | [0.000e+00, 9.191e-06] |

UKB = UK Biobank; AF = allele frequency; CI = confidence interval; NFE = non-Finnish European cohort

Supplementary Table S4 (PDF spreadsheet). Pairwise comparison of the relative risk of carriers of 26 RYR1 variants developing clinical malignant hyperthermia (MH).

RR = relative risk of clinical MH; LR = likelihood statistic; FDR_BH = Benjamini–Hochberg false discovery rate
